# A FAIR layer for the INHERENT haemoglobinopathy patient registry

**DOI:** 10.64898/2026.09.16.26363187

**Authors:** Stella Tamana, Christina Yiangou, Kalia Orphanou, Maria Xenophontos, Panayiota L. Papasavva, César Bernabé, Marco Roos, Daphne Wijnbergen, Martijn G. Kersloot, Ronald Cornet, Anna Minaidou, Coralea Stephanou, Sotiroula Chatzimatthaiou, Annalisa Landi, Viviana Giannuzzi, Fedele Bonifazi, Carsten W. Lederer, Petros Kountouris

## Abstract

Haemoglobinopathy registries support research and outcome monitoring. Still, reuse is limited due to heterogeneous structures, registry-specific coding, and incomplete semantic representation. We designed and implemented a FAIRification workflow for the INHERENT haemoglobinopathy platform, an international genotype-phenotype registry, as part of the HemaFAIR project. The workflow was extended to the Cyprus Haemoglobinopathy Patient Registry to demonstrate its applicability across a second registry. Source data and metadata were transformed through two independent but complementary harmonisation branches executed in parallel: one producing an OMOP CDM representation, and the other generating a CARE-SM representation. The workflow generated graph-based semantic resources, predefined query services, public aggregate dashboards, and application programming interface (API) endpoints. Registry metadata were published through the European Rare Disease Registry Infrastructure and a FAIR Data Point. These outputs support findability, interoperable analysis and controlled reuse while preserving existing governance over patient-level data.

## Introduction

Haemoglobinopathies, including thalassaemia syndromes and sickle cell disease, are chronic, inherited blood disorders that require lifelong monitoring and multidisciplinary care^1,2^. Although some haemoglobinopathies are highly prevalent in specific regions, such as Sub-Saharan Africa, the Mediterranean, and the Middle East, patient populations are often dispersed across countries, centres, and healthcare systems^3^. Patient registries are therefore essential for outcome monitoring, genotype–phenotype research, service planning, and international collaboration^4–7^.

Within an individual registry, the value of these data depends on robust governance and the ability to link clinical, laboratory, treatment, longitudinal and genetic information over time^8^. However, collaborative reuse across registries remains constrained by structural and semantic heterogeneity. Rare disease registries are often implemented as local or project-specific systems, with differences in eligibility criteria, electronic case report forms (eCRFs), coding conventions, data dictionaries, follow-up structures, and data capture tools^4,9,10^. Although structured electronic data-capture systems support consistent data entry, validation rules, longitudinal events, and repeatable instruments, registry data are not inherently Findable, Accessible, Interoperable, and Reusable (FAIR)^11^. Standard registry exports generally provide flat tables or application programming interface (API) payloads in registry-specific structures, while much of the context needed for interpretation and reuse remains embedded in project-specific metadata, documentation, and expert knowledge^12,13^. Addressing these limitations requires complementary structural and semantic mappings: from eCRF variables to standard vocabulary concepts, and from registry metadata to interoperable machine-readable representations^14–16^.

Common data models (CDMs) and semantic standards provide complementary routes for addressing this gap by enabling heterogeneous registry data to be represented in standardised, machine-interpretable forms. Examples relevant to registry harmonisation, interoperability and reuse include the Observational Medical Outcomes Partnership Common Data Model (OMOP CDM)^17^, Fast Healthcare Interoperability Resources (FHIR)^18^ and the Clinical and Registry Entries Semantic Model (CARE-SM)^19^. Previous FAIRification studies have included both *de novo* approaches, in which FAIR principles^11^ are incorporated during the design of data collection systems, and retrospective approaches, in which existing datasets are transformed into more interoperable and machine-readable resources. These studies have demonstrated the importance of explicit metadata, controlled vocabularies, semantic representation, and governed data publication for enhancing interoperability and facilitating data reuse^20–24^. Haemoglobinopathy-specific harmonisation efforts, including the Sickle Cell Disease Ontology (SCDO)^25^, the American Society of Hematology (ASH) Research Collaborative Data Hub^26^, and SickleInAfrica^27^ data standardisation activities, further underscore the value of disease-specific semantic resources alongside broader data models and exchange standards. However, reports describing integrated FAIRification workflows for rare disease registries remain relatively limited, particularly those combining common data model harmonisation, semantic representations, metadata publication and machine-accessible dissemination services within a single operational implementation. This gap is reflected in recent evidence from the European Cooperation in Science and Technology (COST) Action “Haemoglobinopathies in European Liaison of Medicine and Science” (HELIOS)^28^, which indicates that, despite willingness among haemoglobinopathy research centres to share data, the practical adoption of metadata standards, ontologies and CDMs remains limited^29^.

To address these challenges, the Fostering F.A.I.R Data and Standards in Rare Haematological Diseases (HemaFAIR)^30^ project was established to develop practical methods and interoperable resources for improving the FAIRness of rare haematological disease registries. HemaFAIR focuses on harmonising registry data and metadata while preserving existing governance arrangements and supporting reuse through complementary analytical and semantic representations. In this study, we describe a FAIRification workflow, developed within the HemaFAIR project, which transforms heterogeneous source data into harmonised semantic representations and standardised analytical models, and enables controlled access to aggregate information while preserving data provenance and existing governance arrangements. The HemaFAIR workflow was tailored and demonstrated on the platform of the International Hemoglobinopathy Research Network (INHERENT)^31^, an international registry developed to support large, multi-ethnic genotype–phenotype studies of haemoglobinopathies^31,32^, and extended to the Cyprus Haemoglobinopathy Patient Registry (CYHAPR) as an external contributing national registry node. Within this implementation, INHERENT^31,32^ serves as the central registry platform, whereas CYHAPR contributes patient-level data under its own governance framework while retaining its local registry configuration. Together, these components demonstrate a practical workflow for FAIRifying haemoglobinopathy registry data while supporting interoperable analysis, semantic representation, metadata publication and controlled data reuse across registries without disrupting routine registry operations.

## Methods

### Ethics and governance

This work was conducted as part of the EU-funded HemaFAIR^30^ project and involved FAIRifying data and metadata from the INHERENT^32^ haemoglobinopathy patient registry. INHERENT operates under its established governance, ethics requirements, consent and data access requirements^31,33^. Other registries contributing data within the broader HemaFAIR ecosystem remain subject to their own governance, ethical approval, consent and data-access requirements. The manuscript describes a FAIRification workflow and associated infrastructure; it does not report interventional research or newly collected clinical-study data. No personal information or individual-level patient records are disclosed. For this reason, no ethics approval is required for the present study. The FAIRification workflow did not expand access to patient-level data, which remained accessible only to pre-authorised data controllers under the established INHERENT governance framework in compliance with applicable data protection regulations, including the General Data Protection Regulation (GDPR), Reg (EU) 2016/679^34^. Public-facing outputs, including dashboard indicators, Resource Description Framework (RDF)^35^-derived query outputs and metadata descriptions, are limited to metadata and aggregate or machine-readable representations from curated and controlled data layers.

### Registry source and data scope

The FAIRification workflow was implemented with the INHERENT Research Electronic Data Capture (REDCap) registry as the operational source system^12,32^. The registry and its pilot implementation, as well as the structure and design principles of the INHERENT eCRF, have been described previously^32,36^. The public eCRF documentation and data dictionary are available at https://crf.inherentnetwork.org/ (accessed 21 July 2026), while a versioned copy has been deposited in Zenodo^36^.

For this workflow, INHERENT provided structured patient-level data and metadata, including demographics, consent-related information, diagnoses, genotypes and genetic variants, transfusion status, treatments, laboratory measurements, diagnostic imaging and organ assessments, medical and surgical history, chronic complications, and annual follow-up data^36^. Records were included when the age at registration was greater than two years and the diagnosis field was completed^32^. Eligible records were subsequently processed according to the workflow described below (**Figure 1**).

**Figure 1.**
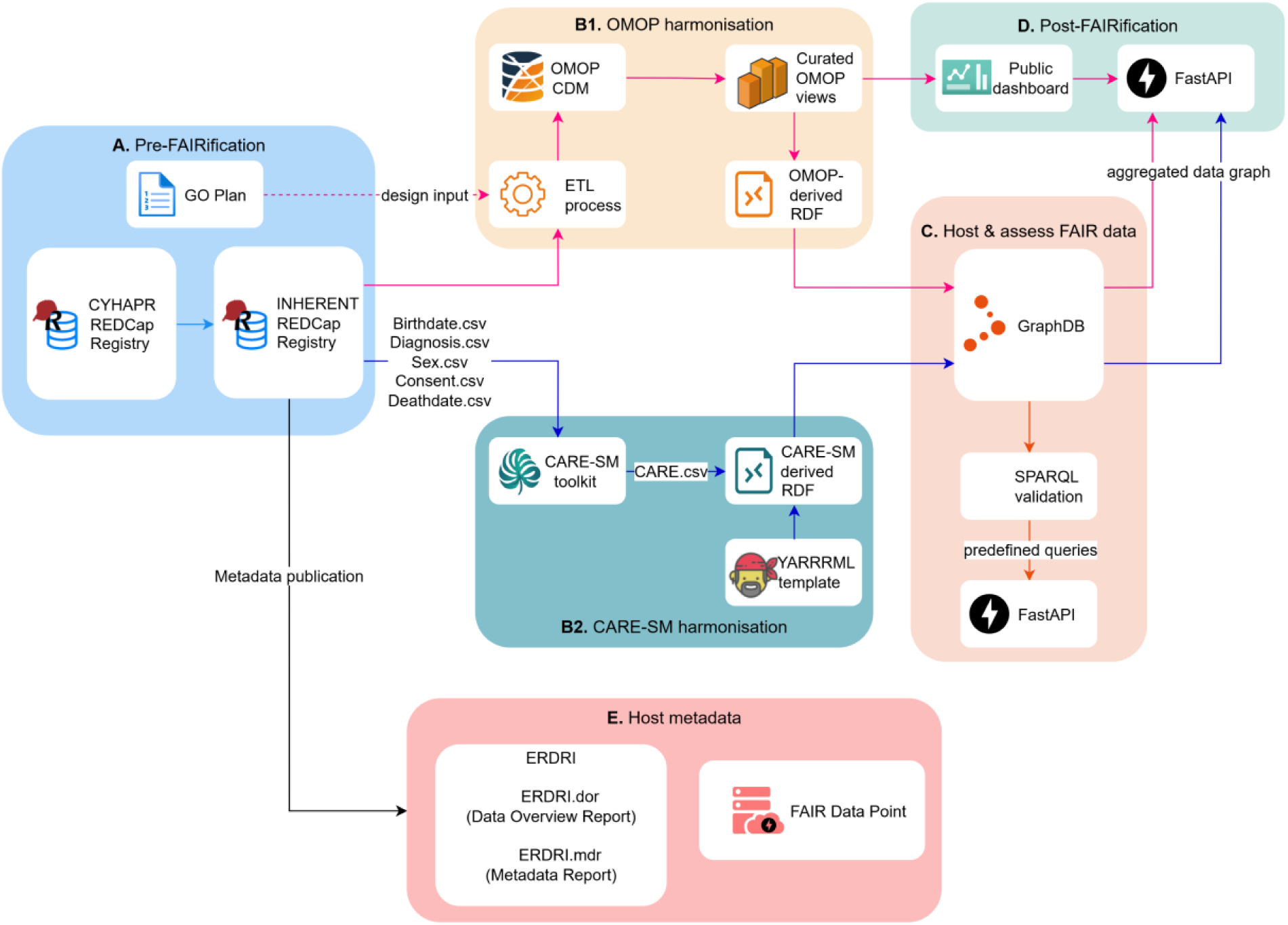
High-level architecture of the HemaFAIR FAIRification workflow. **(A)** Pre-FAIRification planning and preparation of INHERENT and CYHAPR REDCap data and metadata. **(B)** Parallel harmonisation through **(B1)** the OMOP Common Data Model and **(B2)** CARE-SM. **(C)** RDF generation, GraphDB representation and semantic validation using predefined SPARQL queries. **(D)** Controlled public dissemination through dashboards, APIs, structured exports and aggregate RDF resources. **(E)** Metadata publication through ERDRI.dor, ERDRI.mdr and the HemaFAIR FAIR Data Point. Arrow colours distinguish the OMOP, CARE-SM, semantic-validation and metadata-publication pathways.

### FAIR-layer design

The HemaFAIR workflow implements a FAIR layer over the operational INHERENT registry, preserving routine data entry while transforming source data and metadata into harmonised, semantically represented and reusable resources (**Figure 1**). It comprises pre-FAIRification planning and source-metadata preparation (**Figure 1A**), followed by two complementary harmonisation branches executed in parallel: the OMOP CDM branch (**Figure 1B1**) and the CARE-SM branch (**Figure 1B2**). Their outputs support semantic exploration and competency-question validation (**Figure 1C**), controlled public dissemination through dashboards, APIs and machine-readable aggregate resources (**Figure 1D**), and metadata publication through European rare disease infrastructures and a FAIR Data Point (**Figure 1E**).

### Goal-oriented design and competency questions

The design phase was guided by GO-Plan^37^, a goal-orientated method for identifying FAIRification objectives and translating reuse needs into implementation requirements (**Figure 1A**). GO-Plan^37^ was used to define FAIRification objectives, stakeholder roles, reuse scenarios and competency questions (CQs). The full conceptual modelling process is described in the **Supplementary File**, section “Goal-orientated conceptual modelling for the INHERENT FAIRification workflow”.

The CQs were used as implementation and validation targets across the OMOP CDM^17,38^ branch, the RDF^35^ representation, the GraphDB^39^ query environment and the dashboard-facing outputs. They informed the selection of source variables, OMOP^38^ domains, RDF graph patterns, PARQL Protocol and RDF Query Language (SPARQL)^40^ queries and aggregate indicators needed to test whether the workflow could support meaningful reuse scenarios.

### Source eCRF semantic annotation

The HemaFAIR workflow used semantic information from the INHERENT eCRF^36^ as structured metadata input for the OMOP CDM^38^ and CARE-SM^19^ harmonisation branches. This information included ontology codes, source-unit mappings and conversions, and predefined response options. In this context, predefined response options refer to set value lists of fields in REDCap^12^, such as dropdown lists, radio buttons, and checkboxes, where local response codes, including numerical choices, correspond to human-readable labels and require explicit mapping to ontology terms or OMOP CDM^38^ concepts.

Branch-specific field-annotation tags in REDCap were used as structured metadata that could be extracted alongside registry metadata through the REDCap^12^ API. For the OMOP harmonisation branch, the *@HFAIR* tag enabled transformation scripts to parse ontology identifiers, source-unit mappings and conversions, and mappings for predefined response options, thereby supporting mapping enrichment, assignment of standard unit concepts, and OMOP concept lookup. Standard OMOP vocabularies were obtained from the Athena^41^ repository using release v20260227, downloaded 4 March 2026, and loaded into the OMOP CDM environment^38^ to support concept identification. The full list of vocabularies loaded into OMOP, including whether each vocabulary was referenced by the *@HFAIR* mapping layer, is provided in **Supplementary Table S9**. For the CARE-SM^19^ branch, the *@CARE* tag captured the information needed to map selected REDCap variables to CARE-SM classes, temporal attributes, and ontology-based value representations. The CARE-SM branch was applied to selected European Platform on Rare Disease (EU RD) Common Data Elements (CDEs)^42^, a core set of data elements recommended to improve comparability and reuse across rare disease registries. The CARE-SM mapping captured temporal information, including *start* and *end* dates where applicable^19^. Ontology annotations were stored as structured lists linking source data element values to standardised vocabularies and controlled terms. These annotations were processed by the transformation scripts, while ontology selection and mappings requiring clinical or semantic judgment were reviewed by domain experts.

### Harmonisation to CARE-SM

The CARE-SM harmonisation workflow, shown in **Figure 1B2**, uses existing tools, i.e., the CARE-SM toolkit^43^, and mapping languages, i.e., YARRRML^44^, for specific processing tasks. These are integrated and executed within a custom extract, transform, and load (ETL) pipeline implemented in Python and containerised using Docker^45^. As an initial step, we use the *@CARE* annotation tags, as described in the previous section, to align the CDE elements with the CARE-SM^19^ data model. Using this mapping configuration, both metadata (e.g., data dictionaries) and observational data are retrieved programmatically from REDCap via its API^12^. The extracted data are then materialised to multiple structured comma-separated value (CSV) files, each corresponding to a distinct CARE-SM class (e.g., *BirthDate.csv*), following a class-centric data organisation approach consistent with the CARE-SM framework^19,43^.

Subsequently, the generated CSV files are processed using the CARE-SM Toolkit, which performs a validation and curation step before semantic transformation into RDF. In the next step, the YARRRML RDFizer^44^ service is invoked to transform the curated CSV files into RDF triples according to the predefined mappings. The output is serialised into a single N-Quads file, enabling the representation of named graphs for different CARE-SM^19^ classes and contexts. Once generated, the RDF data is automatically imported into a dedicated repository in GraphDB^39^, where it becomes available for querying via the SPARQL^40^ endpoints. A FastAPI^46^ application, with Swagger/OpenAPI documentation^47^, was deployed as an intermediary layer over GraphDB to enable execution of predefined SPARQL queries on the imported CARE-SM RDF data. The source code, configuration files, mapping artefacts, RDF generation resources, GraphDB deployment resources and dashboard components are maintained in the project’s GitHub repository: https://github.com/cing-mgt/hemafair-fairification-workflow.

### Harmonisation to OMOP CDM

Alongside the CARE-SM branch, the OMOP branch provided the primary harmonised relational representation of the registry’s data for analytics, public aggregate views and downstream RDF^35^ generation. Whereas the CARE-SM branch represents selected rare disease CDEs, the OMOP branch processes the complete registry dataset. Source fields are routed into the appropriate OMOP domains or provenance-preserving mapping structures, while fields requiring additional semantic curation are explicitly tracked within the mapping framework.

Structured registry data were transformed into OMOP CDM version 5.4^38^ via a registry-aware ETL workflow. The ETL was implemented as a modular Python workflow with a main entry point and an orchestration layer that coordinated REDCap extraction, metadata processing, mapping-driven transformation, and OMOP loading. The transformed outputs were loaded into a PostgreSQL-backed OMOP database. The transformation used source-level *@HFAIR* annotations alongside version-controlled mapping files. The mapping template records the registry name, REDCap field, OMOP target table and column, transformation logic, OMOP concept identifiers, unit metadata, and mapping status. For fields with predefined response options, the mapping template preserves the original REDCap codes and labels and, where available, links them to curated ontology terms or OMOP concepts^41^. This supports the consistent transformation of coded responses while retaining provenance for the original registry values. The specific mappings to OMOP CDM tables are summarised in the **Supplementary Table S10**.

Before transforming the data according to the OMOP specification, the ETL performs registry-specific data quality and consistency checks, including validation that the cohort carried forward into OMOP aligns with registry inclusion and exclusion criteria, verification of diagnosis field completeness, date parsing, annual update anchoring, and preservation of source provenance. The OMOP harmonisation scripts and mapping artefacts are maintained in the same project GitHub repository^48^.

### OMOP-derived RDF and dashboard outputs

Curated OMOP-derived views were used to generate two downstream outputs: an RDF representation for semantic validation (**Figure 1C**) and a public aggregate dashboard for interactive exploration (**Figure 1D**). The OMOP-derived RDF resources were generated using ontology-based data access (OBDA) mappings with Ontop^49^ and a lightweight HemaFAIR ontology (companion ontology). The Ontop mappings, companion ontology and associated implementation files used for RDF materialisation are publicly available in the project’s GitHub repository^48^, together with workflow documentation, example configuration files and deployment scripts. The HemaFAIR companion ontology was introduced specifically to expose selected entities and relationships from the curated OMOP-derived views as RDF resources suitable for GraphDB querying and validation. It serves as a lightweight semantic representation of the OMOP branch and is distinct from the separately implemented CARE-SM representation.

For the OMOP harmonisation branch, curated OMOP-derived PostgreSQL views were used as the harmonised source layer for RDF generation (**Figure 2A**). Ontop OBDA mappings and the HemaFAIR companion ontology defined the classes, properties, and mapping rules used to materialise RDF from these views. The resulting OMOP-derived RDF graph captured selected registry entities and relationships from the curated OMOP subset summarised in the **Supplementary Table S10**, while preserving source-facing OMOP values and registry-specific identifiers to support provenance-aware validation and interpretation. The principal entities and relationships exposed through the companion ontology are illustrated in **Figure 2B**.

**Figure 2.**
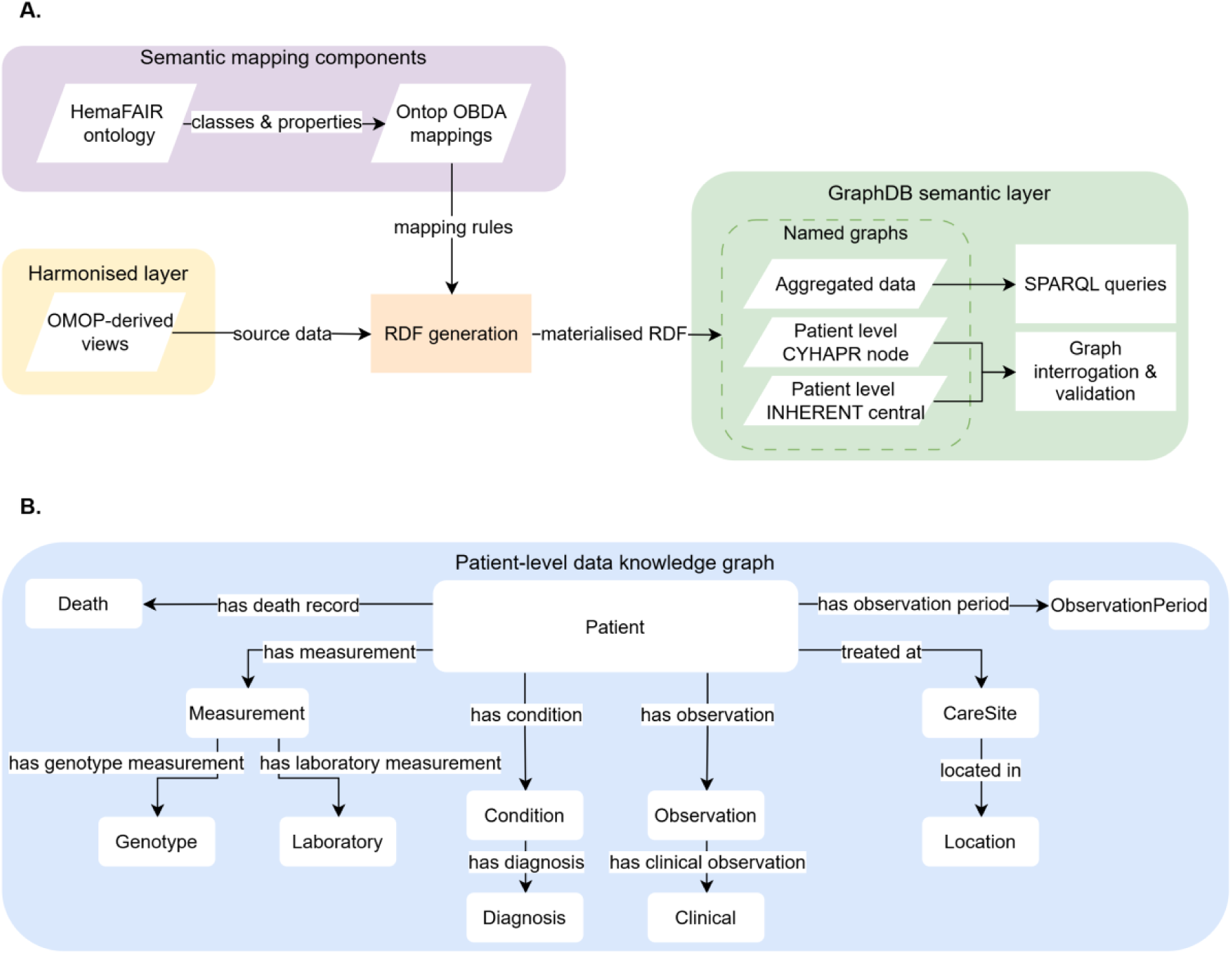
OMOP-derived RDF knowledge graph model and GraphDB workflow. **(A)** Curated OMOP-derived PostgreSQL views, Ontop^49^ OBDA mappings, and the HemaFAIR companion ontology were used to generate materialised RDF. The RDF outputs were loaded into a dedicated GraphDB repository as named graphs for public aggregate data, patient-level CYHAPR data and patient-level INHERENT central-registry data. GraphDB provided the triplestore and query environment for SPARQL querying, graph interrogation and semantic validation. **(B)** Simplified patient-level model of the OMOP-derived RDF represented using the HemaFAIR companion ontology, showing selected registry entities and relationships, including patients, diagnoses, genotype and laboratory measurements, clinical observations, observation periods, death records, care sites and locations.

The resulting RDF resources were loaded into a dedicated GraphDB repository to support graph exploration and SPARQL-based validation against the three predefined CQs listed in the **Supplementary Table S6**. These queries tested whether the RDF representation preserved the relationships required to answer those questions.

The public INHERENT analytics dashboard was implemented as a web application with a React/Vite frontend and a FastAPI^46^ backend. The FastAPI backend provides documented aggregate API endpoints for dashboard indicators, competency-question outputs, and structured exports, using curated OMOP-derived public views rather than patient-level RDF queries. The FastAPI layer is registry-aware: aggregate outputs can be generated from the INHERENT central registry and, for selected CQs and dashboard outputs, can incorporate aggregated contributions from CYHAPR as an external INHERENT node with separate database infrastructure and governance boundaries. This design preserves registry separation while enabling cross-registry aggregate reporting without exposing patient-level records.

### Metadata publication and registry discoverability

To support findability and registry-level discoverability, metadata describing INHERENT were published through the European Rare Disease Registry Infrastructure (ERDRI)^50^ using the Directory of Registries (ERDRI.dor)^51^ and the Metadata Repository (ERDRI.mdr)^52^. A FAIR Data Point (FDP)^53^ was also deployed to expose registry metadata organised into catalogues and datasets, following the official Health Research Infrastructure (Health-RI) metadata model^54^. This model extends the Health-related extension of the Data CATalog Vocabulary Application Profile (HealthDCAT-AP)^55^ and provides more explicit guidance for implementation, supporting alignment with the European Health Data Space (EHDS)^56^ metadata requirements. To enforce metadata compliance across published resources, the Health Research Infrastructure (Health-RI) Shapes Constraint Language (SHACL) profiles were programmatically installed using the FAIRDataPointSchemaTool^54,55^. The FDP is populated and managed programmatically via the FDP API^57^, using the central CRF^36^ as the primary input. Finally, the FDP was assigned a persistent identifier (https://w3id.org/HemaFAIR/fdp) and indexed by the central FAIR Data Point Index^58^.

## Results

We developed and implemented a FAIRification workflow for the INHERENT^32^ haemoglobinopathy central registry (**Figure 1**). The workflow produced five key output layers: (1) a harmonised OMOP CDM^17,38^ representation; (2) RDF^35^ representations derived from both the OMOP and CARE-SM^19^ branches stored in GraphDB^39^; (3) an RDF query layer implemented through two FastAPI^46^ services, each connected to a separate GraphDB repository to execute predefined SPARQL^40^ queries; (4) a public analytics dashboard with structured aggregated export functions; and (5) registry- and dataset-level metadata published through ERDRI.dor^51^, ERDRI.mdr^52^, and FDP^53^. Together, these outputs demonstrate how data and metadata from an operational registry can be transformed into controlled analytical, semantic, aggregate and metadata access layers while preserving source provenance and registry governance boundaries.

### Semantic RDF and GraphDB layer

Both the OMOP and CARE-SM harmonisation branches produced RDF outputs that were imported into dedicated repositories within the same GraphDB instance for semantic exploration and validation (**Figure 1C**). The OMOP-derived RDF graph represented curated registry entities and relationships from the harmonised OMOP layer using a lightweight companion ontology within the HemaFAIR namespace (**Figure 2A**). The CARE-SM-derived RDF graph represented selected EU RD CDEs. Predefined SPARQL queries returned consistent aggregate outputs for the indicators represented in both branches, including total record count, sex distribution, and diagnosis-group counts (**Table 1**).

**Table 1.**
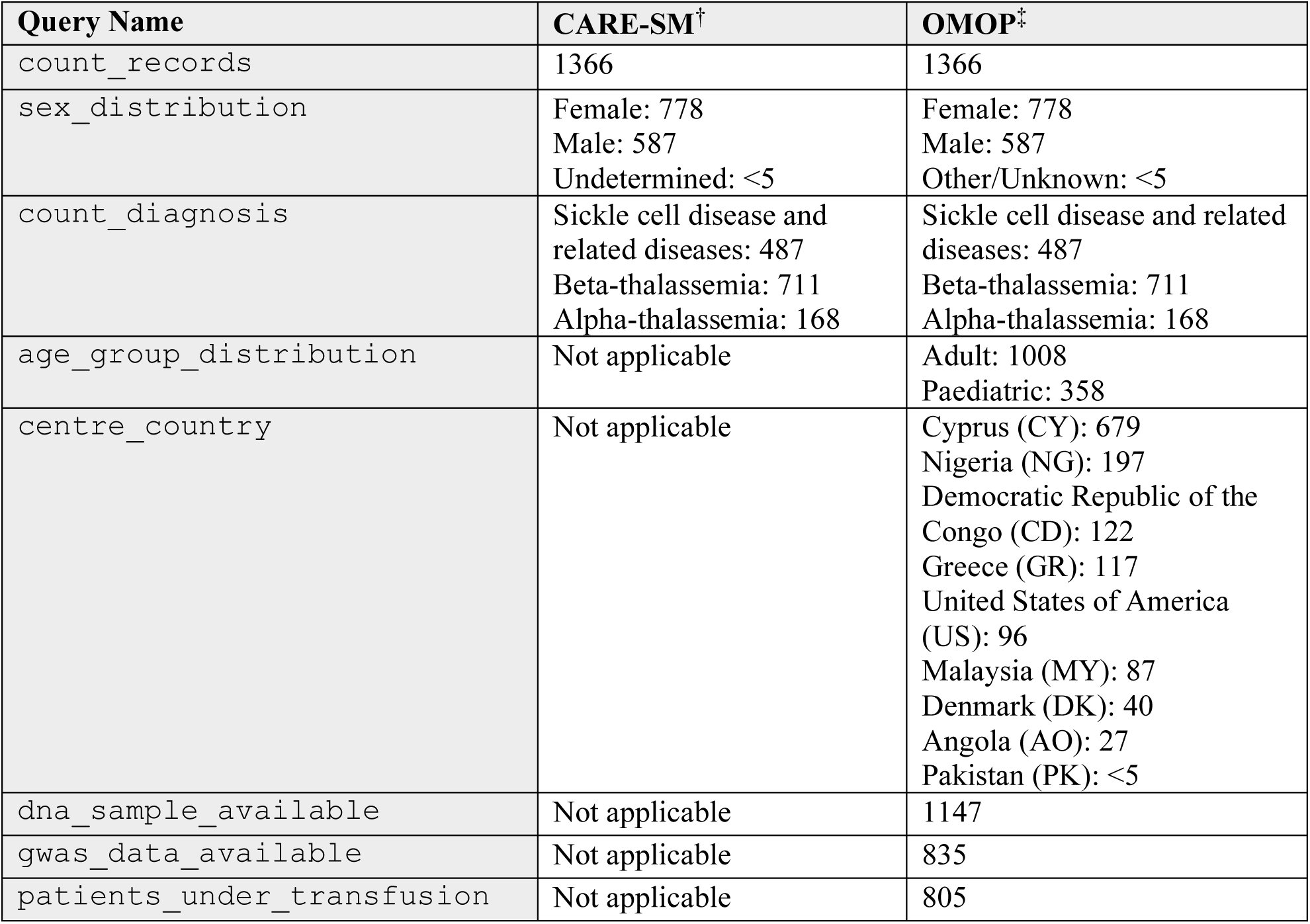
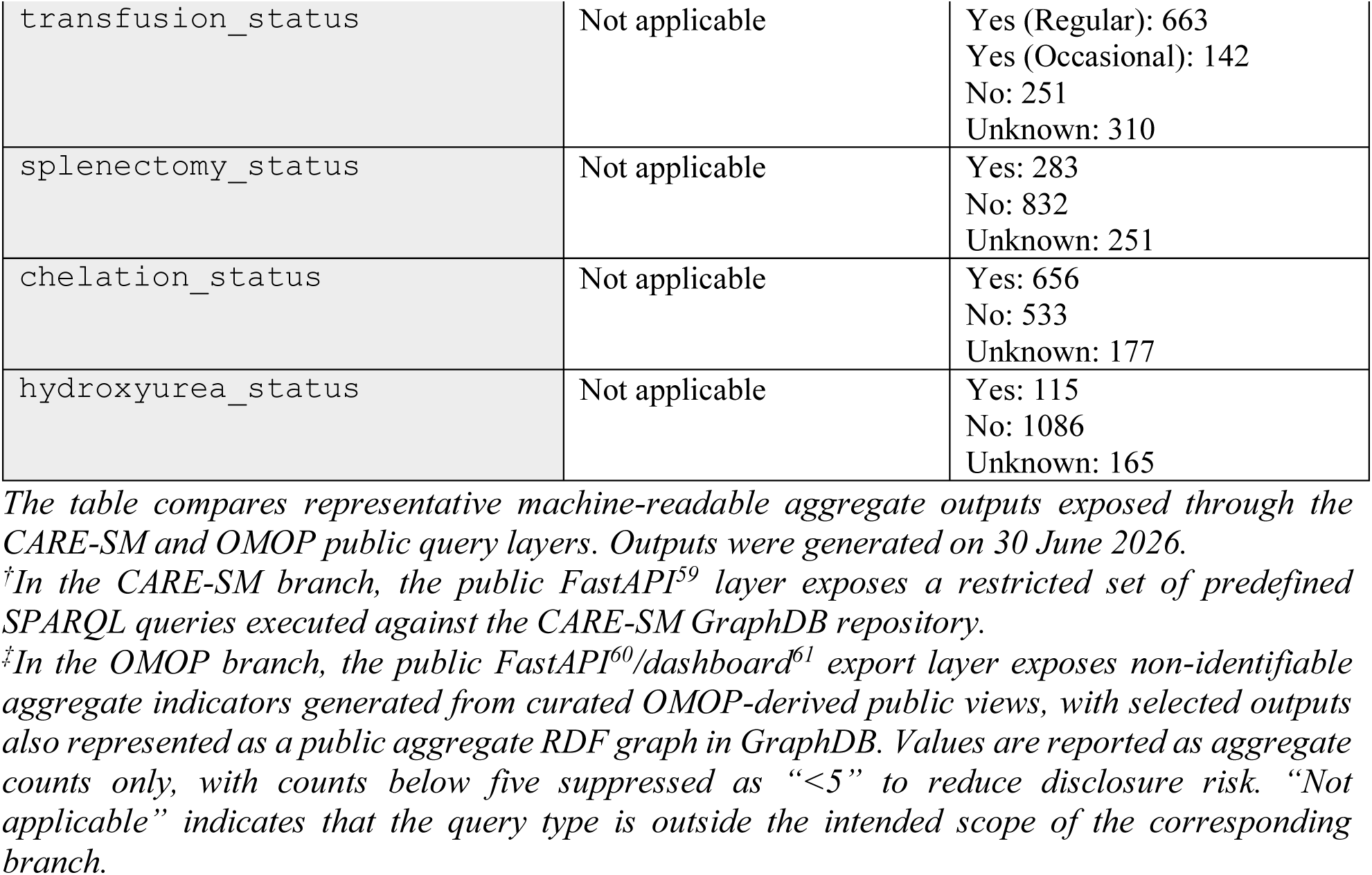
Representative machine-readable aggregate outputs exposed through the public RDF query layers.

Although CARE-SM is designed to be extensible, we deliberately applied it to selected EU RD CDEs, reflecting its original use case and the scope for which it is currently best documented. The broader registry content was harmonised through the OMOP branch, while its lightweight HemaFAIR companion ontology provided an RDF representation of this OMOP-derived content, rather than an independent domain semantic model. The two RDF outputs therefore served complementary purposes: CARE-SM provided a semantically structured representation of the selected CDEs, whereas the OMOP-derived graph supported the broader CQ evaluation and public aggregate outputs.

### Public INHERENT analytics dashboard, APIs, and semantic validation

The HemaFAIR workflow generated a public INHERENT analytics dashboard^61^ that provides interactive access to predefined aggregate registry indicators (**Figure 1D**). The dashboard is available at https://dashboard.inherentnetwork.org/ and uses curated OMOP-derived public views to provide a controlled visual and programmatic access layer for registry-level exploration. In the current implementation, this public output layer is supported by the validated INHERENT and CYHAPR aggregate scope, allowing selected indicators and CQ-orientated summaries to be exposed without releasing patient-level records^61^.

In practice, the dashboard presents headline indicators, including total patient counts, patients with a recorded diagnosis, and selected treatment or procedure indicators such as transfusion status, splenectomy, chelation, and hydroxyurea use^61^. Dynamic filters allow these summaries to be updated by diagnosis group, age group, gender, centre, and country, supporting controlled exploration of registry-level patterns while maintaining the privacy boundary of the underlying registry data^61^. Contextual information panels and popovers embedded throughout the dashboard provide explanatory text, OMOP table and concept links where available, and links to the corresponding public INHERENT eCRF^36^ parameters, making the origin and interpretation of each aggregate indicator transparent to users^61^.

To support programmatic reuse, the same non-identifiable aggregate outputs were made available through documented API endpoints and structured export functions. These included dashboard indicators and CQ outputs, such as CQ1 cohort stratification, CQ2 complication summaries, and CQ3 ferritin-per-complication summaries^60^. The shared export layer enabled the dashboard, manuscript tables and machine-readable aggregate outputs to be generated from the same curated OMOP-derived public views^60^.

SPARQL validation was performed on the OMOP-derived patient-level RDF graph to assess whether it preserved the semantic connections needed to answer the competency questions defined during the GO-Plan^37^ phase. These queries tested cross-domain stratification and aggregation across the OMOP-derived entities represented in the graph. The resulting outputs are presented as evidence of graph connectivity and transformation fidelity rather than as epidemiological analyses. A representative CQ2 SPARQL validation output from the OMOP-derived patient-level RDF graph is shown in **Table 2**. The query was executed against the INHERENT and CYHAPR named graphs in the INHERENT GraphDB repository to test whether the RDF representation preserved the links required to retrieve complication-related records and aggregate them by mapped OMOP condition concept^38,41^. The broader CQ1 stratification output is provided in **Supplementary Table S11,** because of its size and detailed stratification. The full executable SPARQL queries are available in the project repository^48^.

**Table 2.** Representative SPARQL validation output for CQ2: retrieval of recorded iron overload-related and organ-specific complications.

| CQ2<sup>†</sup> complication concept | Condition concept ID | No. patients | Latest annual follow-up year |
| --- | --- | --- | --- |
| Vitamin D deficiency | 436070 | 410 | 2026 |
| Hypogonadism | 4167354 | 128 | 2025 |
| Type 2 diabetes mellitus | 201826 | 125 | 2026 |
| Hypothyroidism | 140673 | 114 | 2025 |
| Atrial fibrillation | 313217 | 69 | 2025 |
| Delayed puberty | 4266651 | 55 | 2024 |
| Cirrhosis of liver | 4064161 | 26 | 2024 |
| Tricuspid valve regurgitation | 4006971 | 25 | 2025 |
| Hypoparathyroidism | 140362 | 23 | 2023 |
| Heart failure | 316139 | 22 | 2023 |
| Sickle cell retinopathy | 4021365 | 20 | 2026 |
| Chronic kidney disease | 46271022 | 14 | 2023 |
| Unprovoked deep vein thrombosis | 46285904 | 8 | 2024 |
| Hypopituitarism | 4254542 | 6 | 2023 |

<sup>†</sup>CQ2 assessed whether complication-related records could be retrieved from the OMOP-derived RDF graph and aggregated by OMOP condition concept. The table lists representative CQ2 complication concepts, their mapped OMOP condition concept identifiers<sup>41</sup>, the number of patients retrieved in the query output, and the latest annual follow-up year represented in those records. Counts below five are suppressed. The year column indicates the annual follow-up context from which records were retrieved and should not be interpreted as the clinical onset or incidence year of complication. Outputs were generated on 30 June 2026.

The representative CQ2 validation output demonstrates that the OMOP-derived RDF graph preserved the links needed to retrieve complication-related records and aggregate them by mapped OMOP condition concept^38,41^ (**Table 2**). The query returned endocrine, metabolic, cardiac, ocular, hepatic, renal, and thrombotic complication concepts, demonstrating that the graph can support cross-domain retrieval across heterogeneous complication categories. These outputs are presented as semantic validation of the RDF representation and transformation logic, rather than as estimates of complication prevalence or incidence. The latest annual follow-up year provides context on the most recent source-derived annual update represented in the query output and should not be interpreted as the clinical onset year of the complication.

The CQ3 ferritin-per-complication summary is presented in **Table 3**. The validation output demonstrated that ferritin measurements could be retrieved from the OMOP-derived measurement layer and summarised across recorded endocrine, cardiac, ocular and hepatic complication groups. Ferritin values are presented as descriptive semantic validation outputs and should not be interpreted as evidence of causal association, clinical risk or epidemiological effect.

**Table 3.** Representative SPARQL validation output for CQ3*^†^* from the OMOP-derived patient-level RDF graphs: ferritin measurements and recorded organ-specific complication groups.

| Organ-specific complication group | No. patients | Min ferritin | Q1 ferritin | Median ferritin | Q3 ferritin | Max ferritin |
| --- | --- | --- | --- | --- | --- | --- |
| Cardiac | 83 | 38 | 385 | 734 | 1835 | 11656 |
| Endocrine | 227 | 36 | 409 | 840 | 2165 | 18523 |
| Hepatic | 20 | 152 | 747 | 1785 | 4638 | 9021 |
| No matched organ-specific complication | 463 | 11 | 312 | 686 | 1515 | 18097 |
| Ocular | 11 | 39 | 164 | 326 | 551 | 2000 |

<sup>†</sup>CQ3 assessed whether ferritin measurements could be retrieved from the OMOP-derived RDF graph and summarised across recorded organ-specific complication groups. The table reports the latest available ferritin measurement per patient within each complication group retrieved from the INHERENT and CYHAPR named graphs. Complication groups may not be mutually exclusive; therefore, patient counts should not be summed across groups. Ferritin values are reported in ng/mL as the minimum, first quartile, median, third quartile and maximum. Values were rounded to the nearest whole number and are presented as descriptive semantic validation outputs rather than evidence of causal association, clinical risk or epidemiological effect. Outputs were generated on 30 June 2026.

Selected outputs from the dashboard and aggregate FastAPI^60^ layer were also represented as machine-readable aggregate RDF resources and loaded into another repository in the same GraphDB instance as CARE-SM and OMOP RDF data. The named graph was used to support SPARQL-based validation and retrieval of the aggregate RDF resources within the GraphDB environment. SPARQL validation identified 113 aggregate observation resources in this graph, comprising 96 CQ1 observations, 13 CQ2 observations, and 4 CQ3 observations. This confirmed that selected public dashboard and aggregate API outputs could be retrieved both through conventional API/export formats and as machine-readable RDF resources.

The metadata publication layer provided complementary registry discoverability outside the dashboard and RDF query environment. INHERENT metadata were represented in ERDRI.dor^51^ and ERDRI.mdr^52^, while the HemaFAIR FDP^53^ exposed machine-readable catalogue, dataset and distribution metadata for discovery and reuse. This separated metadata publication from clinical data access: registry-level and dataset-level descriptions were made openly discoverable, whereas patient-level data remained subject to INHERENT governance and data-access procedures^31,32^.

## Discussion

This work addresses a practical challenge common to rare disease registries: clinically valuable data may be captured in structured electronic systems yet remain difficult to reuse across different research environments. The main contribution of this study is the integration of complementary FAIRification components into a single operational workflow. The implementation combines OMOP CDM^17,38^ and CARE-SM^19^ harmonisation, semantic RDF representations, predefined SPARQL query outputs, metadata publication through ERDRI^51,52^ and the HemaFAIR FDP^53^, public aggregate API and dashboard services, and CQ validation within a reproducible infrastructure^59–61^. By placing a FAIR layer on top of the existing registry, the workflow connects harmonisation, semantic representation, metadata publication and controlled dissemination into an end-to-end process supporting both human and machine reuse, while preventing exposure of patient-level data through the public dissemination layer.

Published FAIRification work has provided both general implementation frameworks and domain-specific solutions for biomedical data. *FAIR in action* provides structured guidance for prospective and retrospective FAIRification across clinical and molecular datasets^62^, while the Swiss Personalised Health Network has demonstrated how Semantic Web technologies, shared RDF schemas and supporting infrastructure can enable standardised representation and interoperable access to health data and metadata^63^. Other initiatives have addressed specific components of this process, including REDCap-to-OMOP transformation^14^, semantic modelling of rare disease CDEs using CARE-SM^19^, and machine-readable metadata publication through FAIR Data Points^64^. This work builds on these complementary developments by integrating source-level semantic preparation, OMOP and CARE-SM transformation branches, RDF materialisation, metadata publication, CQ validation, and controlled public dissemination within an operational multi-registry workflow.

Specifically, we implement a FAIRification workflow supported by parallel independent transformation branches. The OMOP CDM^38^ harmonisation branch was selected to provide a practical foundation for structuring heterogeneous eCRF content into reusable analytical domains. In the INHERENT implementation, OMOP enabled patient-level attributes, diagnoses, measurements, observations, treatments, procedures, care-site information and follow-up periods to be represented in a consistent relational model. This delivered several practical benefits: it separated operational data capture from downstream analytical use; standardised heterogeneous eCRF fields into a common data model; enabled reproducible querying and the generation of curated public views; and preserved source variables, predefined response values and ontology annotations in provenance fields and mapping artefacts. Because the transformation is driven by explicit mapping files, predefined response-option mappings and source-provenance fields, the same approach can be adapted to other registry implementations with different source structures, provided that equivalent source metadata and mapping decisions are available. REDCap2OMOP similarly addresses the practical transformation of evolving REDCap projects into the OMOP CDM^14^, whereas this work places this transformation within a broader FAIRification architecture that also includes semantic RDF representations, registry and dataset metadata publication, public aggregate services and cross-registry CQ validation.

In parallel, the CARE-SM^19^ model was implemented as a knowledge graph-based representation focused on semantic interoperability. Its application in the present workflow was intentionally restricted to EU RD CDEs^42^ because the CDE modules represent the original and currently most extensively documented CARE-SM use case^19^. This provides immediate interoperability value for alignment with rare disease resources, including the European Rare Diseases Research Alliance (ERDERA)^65^ ecosystem. Importantly, CARE-SM is designed to be extended through its core data element semantic pattern. Representing these models in RDF could therefore provide a basis for mapping the lightweight HemaFAIR ontology annotations to additional CARE-SM concepts, supporting future cross-registry data sharing.

The OMOP-derived aggregated outputs were further exposed through a public dashboard and documented FastAPI endpoints^60,61^. This public layer supports controlled exploration of aggregate indicators generated from curated OMOP-derived public views and provides a complementary interactive and programmatic reusability layer^61^. Selected dashboard and FastAPI outputs were also represented as public aggregate RDF resources, ensuring that the same non-identifiable summaries could be reused through visual, API-mediated and machine-readable semantic access routes without exposing individual-level data^60^.

Our workflow also highlights the importance of source-level semantic preparation and curated mapping artefacts. REDCap Field annotations (i.e., @HFAIR and *@CARE*), mapping templates, and value maps can provide structured metadata that can be parsed during transformation and carried through to OMOP and RDF outputs^61^. However, these resources require expert curation and should not be treated as fully automatic semantic annotation. Their value lies in making mapping decisions explicit, version-controlled, and reproducible, rather than in hiding them in transformation code or relying on manual interpretation of exported spreadsheets. Although semantic annotation is ideally incorporated at the point of data capture, mapping resources remain essential for harmonising legacy registries and extending existing systems that were not originally designed with standardised semantic models.

A strength of the HemaFAIR implementation is that the workflow was designed as a reusable, registry-agnostic infrastructure rather than as a one-off transformation specific to INHERENT. The framework was implemented for the INHERENT central registry and for CYHAPR as an external INHERENT node^32^, while preserving registry-specific configuration, database separation, and governance boundaries. This registry-aware design enabled selected public dashboard indicators and CQ-orientated aggregate API outputs to retrieve and combine non-identifiable aggregate results from both platforms where appropriate. It therefore demonstrates the practical benefit of the FAIRification process: additional INHERENT nodes or related haemoglobinopathy registries can be incorporated through shared transformation logic, curated mappings and controlled aggregate output layers. This can be achieved without requiring a fundamental redesign of the registry-to-OMOP transformation, RDF generation, or dashboard API infrastructure. To our knowledge, this represents the first reported end-to-end FAIRification workflow of this scope implemented for haemoglobinopathy patient registries. By enabling registry-specific platforms to contribute through harmonised data structures, reusable semantic mappings and controlled aggregate outputs, while preserving governance separation, the workflow provides a practical foundation for multicentre and multinational collaboration, comparative research and future federated analyses in the haemoglobinopathy field.

### Limitations

Several limitations should be acknowledged. First, the workflow was implemented for the INHERENT central registry and CYHAPR as an external INHERENT national node. Further evaluation across additional INHERENT nodes, haemoglobinopathy registries, sites and eCRF variants will be needed to assess scalability across a wider range of local configurations, governance models and data-completeness profiles.

Second, some semantic extensions are intentionally deferred until further clinical or ontology curation is completed. This includes expanding selected mappings, refining ontology alignment for more complex clinical concepts, and extending the RDF^35^ representation beyond the currently validated entities.

Third, the RDF layer was designed as a controlled semantic validation layer focused primarily on the predefined CQs, rather than as a complete RDF mirror of every transformed REDCap field or OMOP table. This conservative scope improved reproducibility and validation but limited the breadth of SPARQL-queryable content in the initial graph.

Fourth, the public dashboard, FastAPI endpoints, and public aggregate RDF outputs expose only aggregate-level indicators^60,61^. They are intended to support transparent exploration and reuse of non-identifiable summary outputs, but they do not replace governed access to individual-level data for approved research.

Finally, although the workflow is mapping-driven and reproducible, it still depends on expert review of source annotations, value maps and OMOP^38^ concept mappings; these curation steps remain essential for maintaining semantic accuracy across registry versions.

### Future work

Future work will focus on expanding the RDF scope, extending the existing CQ-driven SPARQL query library beyond the current validation examples, and extending FastAPI to support user access controls and parameterised queries across repositories. Further curation of laboratory, treatment, complication and genotype mappings will improve the completeness, consistency and semantic precision of both OMOP-derived outputs, including curated public views, dashboard/FastAPI indicators and RDF representations^61^. Additional work will investigate formal semantic mappings between the lightweight HemaFAIR ontology used for the OMOP-derived RDF and CARE-SM RDF output using established ontology-mapping approaches, with the aim of supporting cross-model reasoning and query interoperability between the two models. Cross-branch CQs will be used to evaluate the resulting semantic integration.

A further priority is to assess how harmonised OMOP data representations can be integrated with RDF outputs and registry-level metadata publication to support federated analysis and learning across INHERENT nodes and related haemoglobinopathy registries. This will require evaluating governance procedures, data-access agreements, cross-node identity management, shared analytical specifications, mapping consistency, and provenance requirements across participating sites. Additional work is also needed to formalise release procedures for versioned code, mapping artefacts, RDF and long-term metadata publication via ERDRI^51,52^ and the HemaFAIR FDP^53^.

## Conclusions

This paper presents a FAIRification workflow developed as part of the HemaFAIR project for the INHERENT^32^ and CYHAPR haemoglobinopathy patient registries, in which registry data and metadata are harmonised through two complementary approaches: (a) the OMOP CDM^38^, which represents registry data in standardised tables and vocabulary concepts to support consistent cohort definition, querying, stratification and comparative analysis, and (b) the CARE-SM^19^, which provides semantic representation of selected European rare disease Common Data Elements and supports alignment with other European rare disease registries. The resulting outputs are exposed through RDF resources, aggregate data services, public dashboard views^61^, metadata records published through ERDRI^51,52^ and the HemaFAIR FDP^53^. Overall, the workflow demonstrates a practical approach to transforming operational rare disease registries, illustrated through its implementation in both the international INHERENT registry and the national CYHAPR registry, irrespective of their underlying data capture systems, into structured, interoperable and reusable data assets. By combining harmonisation, semantic enrichment, metadata publication, predefined query access and dashboard-based dissemination, the workflow provides a reproducible pathway from routine registry data collection to controlled and sustainable data reuse.

## Data Availability

Pseudonymised patient-level data used in the INHERENT registry are not publicly available because they contain sensitive health information and are subject to patient consent, ethical, legal, and registry-governance restrictions. Pseudonymised patient-level data may be shared only where the relevant consent permits reuse and following submission, review and approval of the INHERENT Steering and Data Access Committee. Public-facing outputs generated by the HemaFAIR FAIRification workflow are limited to non-identifiable aggregate indicators, curated registry metadata, public eCRF documentation and machine-readable metadata records. The public INHERENT analytics dashboard is available at https://dashboard.inherentnetwork.org/ and provides non-identifiable aggregate indicators rather than patient-level registry data. Selected aggregate outputs are also exposed through documented API endpoints and machine-readable public aggregate RDF resources, where applicable. The public eCRF documentation is available at https://crf.inherentnetwork.org/ and deposited in Zenodo at https://doi.org/10.5281/zenodo.21260336. Registry-level metadata for INHERENT is available through the European Rare Disease Registry Infrastructure (ERDRI), including the INHERENT registry records in Directory of Registries (ERDRI.dor; https://eu-rd-platform.jrc.ec.europa.eu/erdridor/register/name/INHERENT) and Metadata Repository (ERDRI.mdr; https://eu-rd-platform.jrc.ec.europa.eu/mdr/detail/INHERENT). Dataset-level metadata is also available through the HemaFAIR FAIR Data Point at https://w3id.org/HemaFAIR/fdp. Where CYHAPR contributes to aggregate public outputs as an external INHERENT node, only non-identifiable aggregate indicators are exposed; patient-level CYHAPR data remain subject to the corresponding registry governance and access procedures.

## Code Availability

The source code and documentation for the HemaFAIR FAIRification workflow are publicly available at https://github.com/cing-mgt/hemafair-fairification-workflow. The repository contains implementation materials for both harmonisation branches described in this manuscript: OMOP CDM and CARE-SM. These include workflow documentation, example configuration files, mapping templates, value maps, SQL and SPARQL query examples, Ontop, RDF and GraphDB configuration files, dashboard API and frontend code, CARE-SM workflow files and operational refresh scripts.

The repository supports reproduction and adaptation of the workflow structure but does not contain credentials, local deployment paths, REDCap API tokens, raw registry exports, individual-level registry data, patient-level OMOP data or RDF exports generated from participant data. Package requirements and example execution instructions are provided for use with appropriately authorised source data and local deployment settings.

## Author Contributions

S.T. designed, adapted and implemented the registry-aware FAIRification workflow for the INHERENT use case, including the mapping framework, OMOP CDM harmonisation workflow, OMOP-derived RDF generation, GraphDB loading and validation, dashboard/API layer and aggregate export functions. S.T. performed validation analyses, prepared figures and tables, and drafted the manuscript. C.Y. contributed to REDCap development, eCRF management, registry interpretation, data validation, table and figure preparation, and manuscript revision. K.O. contributed to FAIRification design, competency-question formulation, GO-Plan activities, CARE-SM implementation, semantic modelling, data validation, table and figure preparation, and manuscript revision. M.X. contributed to FAIRification design, competency-question formulation, GO-Plan activities, FAIR Data Point implementation, semantic modelling, metadata publication, data validation, table and figure preparation, and manuscript revision. P.L.P. contributed haematology and registry-domain expertise, including GO-Plan activities, competency-question formulation and clinical interpretation of registry structures. A.L., V.G. and F.B. contributed legal, ethical, governance and dissemination expertise, including review of data-sharing considerations, dissemination strategy and manuscript revision. C.B., M.R., D.W., M.G.K. and R.C. contributed to FAIRification planning, GO-Plan activities, semantic modelling, RDF representation, GraphDB deployment and querying, FAIR data infrastructure, SPARQL validation and manuscript revision. A.M. and C.S. contributed to registry-aware FAIRification planning, CYHAPR registry interpretation, governance considerations, validation and manuscript revision. S.C. contributed to project management, coordination and manuscript revision. C.W.L. provided institutional and departmental oversight and contributed to manuscript revision. P.K. conceptualised the study, secured project funding, supervised the work and revised the manuscript.

All authors reviewed and approved the final manuscript.

## Acknowledgements

The authors gratefully acknowledge the patients and families who contribute data to the INHERENT haemoglobinopathy patient registry, and the clinicians, researchers, data managers, and registry contributors who support data collection, curation, and governance. We also thank members of the HemaFAIR consortium, the HELIOS COST Action, and ERDERA for their input during the FAIRification design process, including discussions on registry interoperability, semantic modelling, metadata publication, and data reuse. This article/publication is based upon work from COST Action HELIOS, CA22119, supported by COST (European Cooperation in Science and Technology).

## References

1. Modell, B. & Darlison, M. Global epidemiology of haemoglobin disorders and derived service indicators. Bull. World Health Organ. 86, 480–487 (2008).

2. Piel, F. B. et al. Defining global strategies to improve outcomes in sickle cell disease: a Lancet Haematology Commission. Lancet Haematol. 10, e633–e686 (2023).

3. Aguilar Martinez, P., et al. Haemoglobinopathies in Europe: health & migration policy perspectives. Orphanet J. Rare Dis. 9, 97 (2014).

4. Bellgard, M. I., Snelling, T. & McGree, J. M. RD-RAP: beyond rare disease patient registries, devising a comprehensive data and analytic framework. Orphanet J. Rare Dis. 14, 176 (2019).

5. Bandeira, M. et al. An overlook on the current registries for rare and complex connective tissue diseases and the future scenario of TogethERN ReCONNET. Front. Med. 9, 889997 (2022).

6. Boulanger, V., Schlemmer, M., Rossov, S., Seebald, A. & Gavin, P. Establishing Patient Registries for Rare Diseases: Rationale and Challenges. Pharm. Med. 34, 185–190 (2020).

7. Baldomero, H. et al. The role of registries in hematological disorders. Best Pract. Res. Clin. Haematol. 37, 101556 (2024).

8. Giannuzzi, V. et al. Ethical and procedural issues for applying researcher-driven multi-national paediatric clinical trials in and outside the European Union: the challenging experience of the DEEP project. BMC Med. Ethics 22, 49 (2021).

9. Atalaia, A. et al. EURO-NMD registry: federated FAIR infrastructure, innovative technologies and concepts of a patient-centred registry for rare neuromuscular disorders. Orphanet J. Rare Dis. 19, 66 (2024).

10. Bassanese, G. et al. The European Rare Kidney Disease Registry (ERKReg): objectives, design and initial results. Orphanet J. Rare Dis. 16, 251 (2021).

11. Wilkinson, M. D. et al. The FAIR Guiding Principles for scientific data management and stewardship. Sci. Data 3, 160018 (2016).

12. Harris, P. A. et al. The REDCap consortium: Building an international community of software platform partners. J. Biomed. Inform. 95, 103208 (2019).

13. Harris, P. A. et al. Research electronic data capture (REDCap)—A metadata-driven methodology and workflow process for providing translational research informatics support. J. Biomed. Inform. 42, 377–381 (2009).

14. Gurley, M. J., Warner, J., Bushmanova, Y. & Wehbe, F. REDCap2OMOP: A platform for ETLing REDCap projects into the OMOP CDM. https://www.ohdsi.org/wp-content/uploads/2021/09/65_poster-REDCap2OMOP.pdf (2021).

15. Cheng, A. C. et al. REDCap on FHIR: Clinical Data Interoperability Services. J. Biomed. Inform. 121, 103871 (2021).

16. Dos Santos Vieira, B., et al. Towards FAIRification of sensitive and fragmented rare disease patient data: challenges and solutions in European reference network registries. Orphanet J. Rare Dis. 17, 436 (2022).

17. Voss, E. A. et al. Feasibility and utility of applications of the common data model to multiple, disparate observational health databases. J. Am. Med. Inform. Assoc. JAMIA 22, 553–564 (2015).

18. Bender, D. & Sartipi, K. HL7 FHIR: An Agile and RESTful approach to healthcare information exchange. in Proceedings of the 26th IEEE International Symposium on Computer-Based Medical Systems 326–331 (2013). doi:10.1109/CBMS.2013.6627810.

19. Kaliyaperumal, R. et al. Semantic modelling of common data elements for rare disease registries, and a prototype workflow for their deployment over registry data. J. Biomed. Semant. 13, 9 (2022).

20. Groenen, K. H. J., Jacobsen, A., Kersloot, M. G. et al. The de novo FAIRification process of a registry for vascular anomalies. Orphanet J. Rare Dis. 16, 376 (2021).

21. Lalout, N. et al. The FAIR journey of a patient-driven registry: Reflections and practical solutions from the Duchenne Data Platform FAIRification experience. J. Neuromuscul. Dis. 22143602251382969 (2025) doi:10.1177/22143602251382969.

22. Sinaci, A. A. et al. From Raw Data to FAIR Data: The FAIRification Workflow for Health Research. Methods Inf. Med. 59, e21–e32 (2020).

23. Arefolov, A. et al. Implementation of the FAIR Data Principles for Exploratory Biomarker Data from Clinical Trials. Data Intell. 3, 631–662 (2021).

24. Kersloot, M. G. et al. De-novo FAIRification via an Electronic Data Capture system by automated transformation of filled electronic Case Report Forms into machine-readable data. J. Biomed. Inform. 122, 103897 (2021).

25. Sickle Cell Disease Ontology Working Group. The Sickle Cell Disease Ontology: enabling universal sickle cell-based knowledge representation. Database 2019, baz118 (2019).

26. Wood, W. A. et al. ASH Research Collaborative: a real-world data infrastructure to support real-world evidence development and learning healthcare systems in hematology. Blood Adv. 5, 5429–5438 (2021).

27. Makani, J. et al. SickleInAfrica. Lancet Haematol. 7, e98–e99 (2020).

28. Chatzimatthaiou, S. et al. HELIOS Action: Advancing research, education, and equity in hemoglobinopathies across Europe and beyond. HemaSphere 9, e70258 (2025).

29. Tamana, S. et al. FAIR data gaps and collaboration willingness among hemoglobinopathy research centers. Sci. Data 13, 582 (2026).

30. HemaFAIR. https://hemafairproject.eu/ (2024).

31. Kountouris, P. et al. The International Hemoglobinopathy Research Network (INHERENT): An international initiative to study the role of genetic modifiers in hemoglobinopathies. Am. J. Hematol. 96, E416–E420 (2021).

32. Kountouris, P. et al. Pilot of the International Hemoglobinopathy Research Network for a multiethnic genome-wide association study. Blood Glob. Hematol. 2, 100098 (2026).

33. Didio, A. et al. Ethical and regulatory requirements for conducting researcher-driven large-scale multinational genetic haematological studies: the INHERENT experience. Health Res. Policy Syst. 23, 101 (2025).

34. Regulation (EU) 2016/679 of the European Parliament and of the Council of 27 April 2016 on the protection of natural persons with regard to the processing of personal data and on the free movement of such data, and repealing Directive 95/46/EC (General Data Protection Regulation). Off. J. Eur. Union L 119, 1–88 (2016).

35. RDF - Semantic Web Standards. https://www.w3.org/RDF/.

36. Archer, N., et al. Multi-Ethnic, Resource-Adaptable Case Report Form (CRF) by the International Hemoglobinopathy Research Network (INHERENT). Zenodo 10.5281/zenodo.21260336 (2026).

37. Bernabé, C. et al. GO-Plan: A goal-oriented method for FAIRification planning. Inf. Serv. Use 45, 100–124 (2025).

38. OMOP CDM v5.4. https://ohdsi.github.io/CommonDataModel/cdm54.html.

39. What is GraphDB? — GraphDB 11.3 documentation. https://graphdb.ontotext.com/documentation/11.3/.

40. Polleres, A. SPARQL. in Encyclopedia of Social Network Analysis and Mining 1960–1966 (Springer, New York, NY, 2014). doi:10.1007/978-1-4614-6170-8_124.

41. Odysseus Data Services Inc. Athena – OHDSI Vocabularies Repository. https://athena.ohdsi.org/search-terms/start (2025).

42. European Platform on Rare Disease EU-RD. https://eu-rd-platform.jrc.ec.europa.eu.

43. Alarcón Moreno, P. & Aguiló Castillo, S. CARE-SM Toolkit. GitHub https://github.com/CARE-SM/CARE-SM-Toolkit (2026).

44. Van Assche, D., Delva, T., Heyvaert, P., De Meester, B. & Dimou, A. Towards a more human-friendly knowledge graph generation & publication. in Proceedings of the ISWC 2021 Posters, Demos and Industry Tracks: From Novel Ideas to Industrial Practice vol. 2980 (CEUR-WS.org, 2021).

45. Merkel, D. Docker: lightweight Linux containers for consistent development and deployment. Linux J. 2014, 2:2 (2014).

46. Ramírez, S. FastAPI. https://fastapi.tiangolo.com.

47. OpenAPI Specification - Version 3.1.0 | Swagger. https://swagger.io/specification/.

48. HemaFAIR. HemaFAIR FAIRification Workflow. GitHub https://github.com/cing-mgt/hemafair-fairification-workflow.

49. Xiao, G. et al. The Virtual Knowledge Graph System Ontop. in The Semantic Web – ISWC 2020 (eds Pan, J. Z. et al.) 259–277 (Springer International Publishing, Cham, 2020). doi:10.1007/978-3-030-62466-8_17.

50. European Platform on Rare Disease Registration. https://eu-rd-platform.jrc.ec.europa.eu.

51. ERDRI.dor - International Hemoglobinopathy Research Network. European Rare Disease Registry Infrastructure https://eu-rd-platform.jrc.ec.europa.eu/erdridor/register/name/INHERENT.

52. ERDRI.mdr - International Hemoglobinopathy Research Network. European Rare Disease Registry Infrastructure https://eu-rd-platform.jrc.ec.europa.eu/mdr/detail/INHERENT.

53. FAIR Data Point. INHERENT https://w3id.org/HemaFAIR/fdp.

54. Stichting Health-RI. Health-RI Core Metadata Schema. (2025).

55. HealthDCAT AP | European Health Information Portal. https://www.healthinformationportal.eu/healthdcat-ap.

56. Hussein, R., Gyrard, A., Abedian, S., Gribbon, P. & Martínez, S. A. Interoperability Framework of the European Health Data Space for the Secondary Use of Data: Interactive European Interoperability Framework–Based Standards Compliance Toolkit for AI-Driven Projects. J. Med. Internet Res. 27, e69813 (2025).

57. Benhamed, O. M. et al. The FAIR Data Point: Interfaces and Tooling. Data Intell. 5, 184–201 (2023).

58. daSilvaSantos, L., Burger, K., Kaliyaperumal, R. & Wilkinson, M. FAIR Data Point: A FAIR-Oriented Approach for Metadata Publication. Data Intell. 5, 163–183 (2023).

59. Semantic Query API for INHERENT Registry Using CARE-SM Harmonization - Swagger UI. https://fastapi.inherentnetwork.org/docs.

60. HemaFAIR - INHERENT. INHERENT Dashboard API - Swagger UI. *INHERENT* https://dashboard.inherentnetwork.org/inherent-dashboard-api/docs.

61. INHERENT. INHERENT Dashboard. INHERENT Public Dashboard https://dashboard.inherentnetwork.org/.

62. Welter, D. et al. FAIR in action - a flexible framework to guide FAIRification. Sci. Data 10, 291 (2023).

63. Touré, V. et al. FAIRification of health-related data using semantic web technologies in the Swiss Personalized Health Network. Sci. Data 10, 127 (2023).

64. Wijnbergen, D. et al. The FAIR data point populator: collaborative FAIRification and population of FAIR data points. BMC Med. Inform. Decis. Mak. 25, 211 (2025).

65. European Rare Diseases Research Alliance. ERDERA https://erdera.org/.

