## Supplementary File for "A FAIR layer for the INHERENT haemoglobinopathy patient registry"

Stella Tamana *et al.*

### Goal-oriented FAIRification planning and conceptual modelling for the HemaFAIR workflow

#### Purpose of this supplementary file

This supplementary file outlines the goal-oriented FAIRification planning and conceptual modelling process that guided the HemaFAIR workflow for the International Hemoglobinopathy Research Network (INHERENT)[1,2] haemoglobinopathy patient registry. Adapted from GO-Plan[3], the process extended beyond the conceptual definition of data entities and relationships to define intended reuse scenarios, stakeholders, semantic scope, competency questions (CQs) and implementation requirements. These outputs informed the mapping-driven Observational Medical Outcomes Partnership Common Data Model (OMOP CDM)[4,5] transformation, RDF[6] materialisation, GraphDB[7] loading and querying, dashboard outputs and metadata publication[8–10].

The supplementary material documents the design rationale for the Findable, Accessible, Interoperable and Reusable (FAIR)[11] layer described in the main manuscript. It does not present additional clinical findings. Instead, it records how the FAIRification objectives were translated into implementation requirements for the INHERENT registry and for its registry-aware extension to the Cyprus Haemoglobinopathy Patient Registry (CYHAPR) as an external INHERENT node.

#### Project goals

The FAIRification process was embedded within HemaFAIR[12], a project focused on improving the reuse, interoperability and machine-actionability of rare haematological disease data. In this context, the INHERENT registry was selected as the primary implementation case because it is an operational haemoglobinopathy registry that captures patient-level clinical, longitudinal and genotype-related data through Research Electronic Data Capture (REDCap)[13].

The primary goals of the FAIRification workflow were to:

1. transform operational INHERENT registry data and metadata into a harmonised OMOP CDM[5] layer;
2. preserve source provenance, including source variable names, coded values, ontology annotations, and curated mapping decisions;
3. support semantic representation through RDF materialisation and GraphDB-based querying, using a controlled subset of curated OMOP-derived outputs;
4. enable aggregate public visualisation through curated OMOP-derived dashboard views;
5. expose selected non-identifiable aggregate outputs through documented FastAPI endpoints[14] and machine-readable public aggregate RDF resources;
6. expose registry- and dataset-level metadata through European Rare Disease Registry Infrastructure (ERDRI)[15] and a FAIR Data Point (FDP)[16];
7. provide a reproducible workflow that can be refreshed as the INHERENT registry evolves;
8. support CQ-driven validation of the FAIR layer.

CYHAPR is part of the broader HemaFAIR haemoglobinopathy registry ecosystem and contributes patient-level registry data to INHERENT as an external INHERENT node[1,2]. This contribution informed registry-aware design considerations, including configuration flexibility, database separation, aggregate integration and governance separation. The conceptual modelling and implementation outputs described in the main manuscript remain centred on the INHERENT FAIRification use case, with CYHAPR used to demonstrate registry-aware reuse and controlled aggregate contribution.

#### Stakeholder groups

The FAIRification workflow involved several stakeholder groups with complementary roles (**Supplementary Table S1**). These roles denote areas of expertise rather than mutually exclusive individuals; in practice, the same person or organisation may contribute to more than one role during the FAIRification process.

**Supplementary Table S1. Stakeholder groups involved in the INHERENT FAIRification workflow.**

| Stakeholder group | Role[1] in the FAIRification workflow |
|---|---|
| **Clinical data providers and registry staff** | Enter, validate and maintain patient-level registry data in REDCap[13]. |

| | |
|---|---|
| **Haematologists and clinical experts** | Define clinically meaningful data domains, validate CQs and interpret registry outputs. |
| **Molecular genetics experts** | Support interpretation of genotype, gene and variant-related data elements. |
| **Biocurators and data stewards** | Support terminology curation, mapping validation, data-quality review and provenance preservation. |
| **FAIRification specialists** | Coordinate the FAIRification workflow and ensure alignment with FAIR principles[11]. |
| **Semantic modellers and ontology experts** | Define semantic types, ontology links, RDF structures and graph-query requirements. |
| **OMOP and ETL developers** | Implement REDCap extraction, mapping-driven transformation and OMOP loading. |
| **RDF/SPARQL and GraphDB experts** | Support RDF materialisation, graph loading and SPARQL validation. |
| **Metadata specialists** | Support ERDRI metadata deposition and FAIR Data Point publication. |
| **Dashboard and software developers** | Implement public aggregate visualisation and dashboard, API and frontend deployment. |
| **Reuse stakeholders** | Researchers, clinicians, registry coordinators, policy stakeholders and patient organisations interested in aggregate registry outputs and FAIRified resources. |

[1] *Roles represent areas of expertise and responsibility rather than mutually exclusive stakeholder categories; the same individual or organisation may contribute to more than one role.*

#### Resources included in the FAIRification workflow

The FAIRification workflow drew on several source resources, including mapping artefacts and downstream output layers (**Supplementary Table S2**). These resources represent the practical components considered during GO-Plan conceptual modelling and implementation alignment. They include the primary INHERENT registry resources, CYHAPR as an external INHERENT node used to test registry-aware reuse, curated semantic mapping artefacts, harmonised OMOP outputs, RDF and GraphDB resources, public dashboard views, and metadata publication layers.

**Supplementary Table S2. Resources included in the HemaFAIR FAIRification workflow for the INHERENT use case.**

| Resource | Description | FAIRification role |
|---|---|---|
| **INHERENT REDCap registry** | Operational REDCap-based haemoglobinopathy registry containing patient-level clinical, longitudinal and genotype-related data. | Primary source system for the FAIRification workflow. |
| **INHERENT eCRF and data dictionary** | Structured registry metadata, including variables, coded choices, forms, repeatable instruments and field annotations. | Basis for semantic annotation, mapping generation and metadata-driven transformation of registry data. |

| Resource | Description | FAIRification role |
|---|---|---|
| **CYHAPR REDCap registry** | External INHERENT node containing haemoglobinopathy registry data captured through a REDCap implementation aligned with the INHERENT data model. | Supports validation of the registry-aware workflow by demonstrating adaptation to an external node, preservation of registry-specific configuration, and controlled contribution to aggregate public outputs. |
| ***@HFAIR* field annotations** | Structured REDCap annotations containing ontology references, unit metadata and predefined response-option mappings where applicable. | Used as executable metadata during mapping-driven registry-to-OMOP transformation. |
| **Mapping template and value maps** | Version-controlled mapping artefacts linking REDCap fields and coded values to OMOP tables, columns and concepts. | Serve as the curated semantic contract for ETL execution. |
| **OMOP CDM database** | PostgreSQL-backed harmonised relational layer implementing OMOP CDM v5.4. | Provides the analytical foundation for public views, RDF generation and dashboard indicators. |
| **RDF and GraphDB resources** | Ontop-based RDF materialisation from selected OMOP-derived outputs, with RDF loaded into GraphDB named graphs. | Provides a machine-actionable RDF representation and a GraphDB-based route for SPARQL querying and validation. |
| **Public dashboard views** | Curated OMOP-derived aggregate views consumed by the public INHERENT dashboard. | Provide human-readable aggregate visualisation while avoiding exposure of raw registry data. |
| **ERDRI and FAIR Data Point metadata** | Registry- and dataset-level metadata. | Support findability and machine-readable metadata publication. |

***Note:*** *Resources include source registries, semantic mapping artefacts and downstream FAIR outputs considered during FAIRification planning and implementation.*

#### Domain description

The INHERENT FAIRification domain comprises structured, patient-level haemoglobinopathy registry data captured through an operational electronic registry system[2,17]. The registry includes data relevant to genotype–phenotype research, longitudinal outcome monitoring and aggregate cohort characterisation. Key data domains include demographics, geography, diagnosis, method of diagnosis, genotype and genetic variation, transfusion status, treatments, laboratory measurements, medical and surgical history, chronic complications and annual follow-up information[17].

The FAIRification workflow was designed to transform these operational registry data into reusable outputs without disrupting routine registry data entry. This was achieved by combining source-level semantic annotation, mapping-driven OMOP transformation, RDF materialisation, GraphDB-based validation, public dashboard views and metadata publication.

Semantic precision was supported by using established ontologies and controlled vocabularies where applicable, including Orphanet rare disease nomenclature (ORPHA)[18] for rare disease classification, Systematized Nomenclature of Medicine — Clinical Terms (SNOMED CT)[19] and Human Phenotype Ontology (HPO)[20] for clinical and phenotypic concepts, Logical Observation Identifiers Names and Codes (LOINC)[21] for laboratory measurements, HUGO Gene Nomenclature Committee (HGNC)[22] and Human Genome Variation Society (HGVS)[23] for gene and variant representation, and RxNorm[24] for medicines. The workflow also preserved source-level metadata and coded values, enabling transformed outputs to be traced back to the original eCRF structure.

#### Semantic types

The semantic types below were identified during conceptual modelling to capture the main categories of registry information relevant to FAIRification (**Supplementary Table S3**). They were used to guide OMOP domain routing, RDF modelling and CQ validation. The listed FAIR-layer representations describe the implemented representation where available, or the intended/provenance representation where full semantic modelling was outside the initial RDF scope.

**Supplementary Table S3. Semantic types identified during conceptual modelling.**

| Semantic type | Working definition | Main FAIR layer[1] representation |
|---|---|---|
| **Patient** | Individual enrolled in the INHERENT registry. | OMOP `person`; RDF `Patient`. |
| **Consent** | Recorded permission or restriction related to use or sharing of registry data. | OMOP `observation`; source provenance fields. |
| **Country** | Country of birth, residence or centre location, depending on source context. | OMOP `location`; dashboard filter; RDF `Location`. |
| **Centre** | Healthcare provider institution or registry centre associated with patient care or data contribution. | OMOP `care_site`; dashboard centre distribution; RDF `CareSite`. |
| **Ethnic group** | Self-reported ancestry or ethnicity captured in the registry. | OMOP source/provenance fields; `observation` where applicable. |
| **Diagnosis** | Coded haemoglobinopathy diagnosis. | OMOP `condition_occurrence`; RDF `DiagnosisCondition`. |
| **Method of diagnosis** | Method used to establish or support diagnosis. | OMOP `observation`; mapped source values. |
| **Genotype** | Patient-level disease-related genotype profile. | OMOP `measurement`; RDF `GenotypeMeasurement`. |

| Semantic type | Working definition | Main FAIR layer[1] representation |
|---|---|---|
| **Genetic variant** | Specific gene or variant information, including HGVS/HGNC-related representation where available. | OMOP `measurement`; source provenance and variant lookup artefacts. |
| **Transfusion status** | Registry-derived information on transfusion burden or status. | OMOP `observation`; dashboard indicator/filter. |
| **Laboratory measurement** | Quantitative or coded laboratory result relevant to haemoglobinopathy monitoring. | OMOP `measurement`; RDF `LabMeasurement`. |
| **Clinical complication** | Clinical manifestation, comorbidity or complication recorded in the eCRF. | OMOP `condition_occurrence` or `observation`, depending on mapping decision; RDF `condition/observation` representation where implemented. |
| **Treatment or medication** | Pharmacological or treatment-related intervention. | OMOP `drug_exposure` or `observation`, depending on source field and mapping decision. |
| **Procedure** | Procedure-related information, including selected transfusion-related, surgical or other procedure data. | OMOP `procedure_occurrence`. |
| **Follow-up period** | Annual update or registry follow-up time context. | OMOP `observation_period`; date fields; RDF temporal relationships. |
| **Death or mortality status** | Mortality information where present. | OMOP `death`; RDF `Death`. |

**Note:** *The table lists the main information categories used to guide OMOP domain routing, RDF modelling and competency-question validation.*
*[1] FAIR-layer representations indicate the implemented representation where available, or the intended provenance representation where full semantic modelling was outside the initial RDF scope.*

#### Conceptual relationships

The conceptual model used the CQs to identify relationships that needed to be represented or preserved within the FAIR layer. These relationships informed OMOP domain routing, RDF modelling and validation-query design. The most relevant relationships are summarised in **Supplementary Table S4**.

**Supplementary Table S4. Conceptual relationships represented or preserved in the FAIR layer.**

| Relationship[1] | Description |
|---|---|
| **Patient has diagnosis** | Links a patient to a haemoglobinopathy diagnosis or diagnosis-related condition record. |
| **Patient has genotype** | Links a patient to genotype-related measurements or variant information. |

| Relationship[1] | Description |
|---|---|
| **Genotype supports diagnosis** | Represents the relationship between genotype information and disease classification where this can be inferred from mapped genotype and diagnosis records. |
| **Patient has transfusion status** | Links a patient to transfusion status or transfusion-burden information. |
| **Patient treated at centre** | Links a patient to a care site or contributing centre. |
| **Centre located in country** | Links care-site information to geographic context. |
| **Patient has country of birth or residence** | Captures patient-level geographic context where available. |
| **Patient has laboratory measurement** | Links a patient to temporally indexed laboratory results. |
| **Patient has clinical complication** | Links a patient to complications, comorbidities or complication-related observations. |
| **Patient has treatment or medication** | Links a patient to pharmacological treatment or medication-related records. |
| **Patient has procedure** | Links a patient to procedure-related records, including selected surgical or transfusion-related procedures where mapped. |
| **Patient has follow-up period** | Links a patient to annual update or observation-period context. |
| **Patient has mortality information** | Links a patient to death-related information where available. |

[1] *These relationships were derived from the competency questions and informed OMOP mapping decisions, RDF knowledge-graph modelling and GraphDB-based validation queries.*

These relationships informed both OMOP domain routing and the RDF knowledge graph model. The RDF implementation used a controlled subset of these relationships for the initial materialisation, focusing on patient context, diagnosis-bearing conditions, genotype and laboratory measurements, selected clinical observations, care sites, locations, observation periods, and death records where populated.

#### Reuse stakeholders and intended reuse scenarios

The intended reuse scenarios were identified to ensure the FAIRification workflow supported both registry-specific and broader research needs (**Supplementary Table S5**). These scenarios informed the selection of CQs and the need for multiple reuse layers, including OMOP, RDF knowledge graphs, GraphDB querying, dashboard views, and metadata publication.

**Supplementary Table S5. Reuse stakeholders and intended reuse scenarios.**

| Reuse stakeholder | Intended reuse scenario[1] |
|---|---|
| **Haematologists and clinical researchers** | Cohort characterisation, genotype–phenotype analysis, transfusion-status stratification and complication monitoring. |
| **Registry coordinators and data stewards** | Data-quality review, mapping validation, metadata maintenance and governance-aligned reuse. |

| Reuse stakeholder | Intended reuse scenario[1] |
|---|---|
| **Molecular genetics researchers** | Genotype and variant interpretation in relation to diagnosis and clinical outcomes. |
| **Public health and policy stakeholders** | Aggregate monitoring of registry indicators, disease burden and service-planning metrics. |
| **Patient organisations** | Use of aggregate registry indicators to support communication, advocacy and prioritisation of care needs. |
| **FAIR and semantic-web researchers** | Evaluation of registry-to-OMOP harmonisation, RDF knowledge-graph generation, GraphDB-based querying and metadata publication workflows. |
| **Federated research initiatives** | Future reuse of harmonised and semantically enriched registry outputs in controlled analytical environments. |

[1] *Reuse scenarios informed the selection of competency questions and the design of multiple reuse layers, including OMOP, RDF knowledge graphs, GraphDB-based querying, dashboard views and metadata publication.*

These reuse scenarios informed the selection of CQs and the decision to support multiple reuse layers: OMOP for relational harmonisation, RDF knowledge graphs for semantic representation, GraphDB for SPARQL querying and validation, dashboard views for aggregate visualisation, and ERDRI[25,26]/FDP[16] for metadata discovery.

#### Competency questions

The CQs guided the FAIRification design and defined the minimum set of relationships that should be represented or preserved across the OMOP, RDF knowledge graph, GraphDB query and dashboard-facing layers (**Supplementary Table S6**). CQ1 was selected as the first end-to-end validation case because it links diagnosis, genotype, transfusion status and patient stratification variables. Following broader eCRF implementation in OMOP, CQ2 and CQ3 were retained as additional validation targets for complication and laboratory measurement retrieval.

**Supplementary Table S6. Competency questions guiding the FAIRification workflow.**

| CQ ID | Competency question | Related stakeholders | Data domains involved |
|---|---|---|---|
| **CQ1** | What is the prevalence of specific haemoglobinopathy genotypes and their association with transfusion status across centres, regions, countries or ethnic groups? | Haematologists, registry coordinators, data analysts, public health stakeholders | Diagnosis, genotype, transfusion status, centre, region, country, ethnic group |
| **CQ2** | Which iron overload-related and organ-specific complications are recorded in the registry, and how can these records be retrieved and aggregated by mapped condition concept? | Clinicians, data analysts, registry coordinators | Clinical complications, mapped condition concepts, follow-up period |

| CQ ID | Competency question | Related stakeholders | Data domains involved |
|---|---|---|---|
| **CQ3** | What is the relationship between serum ferritin levels and organ-specific complications? | Haematologists, endocrinologists, cardiologists, clinical researchers | Ferritin measurements, hepatic, cardiac and endocrine complications, longitudinal data |

**Note:** *The competency questions informed the FAIRification workflow design and the validation examples implemented across the OMOP, RDF knowledge-graph, GraphDB-based querying and dashboard-facing layers.*

These CQs informed both the conceptual model and the implemented validation workflow. CQ1 tested end-to-end stratification across diagnosis, genotype, transfusion and geographic or centre-level variables. CQ2 tested whether complication-related records could be retrieved and aggregated by mapped OMOP condition concept. CQ3 tested whether ferritin measurements could be linked to selected complication groups. Together, these questions guided OMOP domain routing, RDF knowledge-graph modelling, loading into GraphDB, SPARQL validation and dashboard-facing aggregate outputs[8,9].

#### Goals emerging from competency questions

The CQs were translated into FAIRification goals to define the implementation requirements for the OMOP, the RDF knowledge graph, and the GraphDB query and dashboard-facing layers (**Supplementary Table S7**). These goals describe the data relationships and transformation capabilities required to answer each question; they should not be interpreted as clinical conclusions.

**Supplementary Table S7. FAIRification goals derived from competency questions.**

| Competency question | FAIRification goal[1] |
|---|---|
| **CQ1** | Represent diagnosis, genotype, transfusion status and patient stratification variables in a way that supports linked querying across OMOP outputs, RDF knowledge graphs, GraphDB-based queries and dashboard-facing outputs. |
| **CQ2** | Represent complication-related fields, mapped condition concepts and follow-up context in a way that supports retrieval and aggregation of recorded iron overload-related and organ-specific complications. |
| **CQ3** | Enable linkage between laboratory measurements and organ-specific complications, supporting temporally contextualised biomarker-to-complication analyses. |

[1] *The goals outline implementation requirements arising from the competency questions rather than clinical conclusions.*

These goals informed decisions on mapping, ontology use, RDF scope definition, SPARQL validation queries and dashboard indicator selection. They also highlighted the need to preserve temporal context, source provenance and mapped concept identifiers throughout the FAIRification workflow.

#### Implementation alignment

The conceptual modelling process informed the following implementation choices (**Supplementary Table S8**). These choices link the GO-Plan requirements to concrete components of the HemaFAIR FAIRification workflow.

**Supplementary Table S8. Alignment between conceptual modelling requirements and implementation decisions.**

| Design requirement | Implementation decision |
|---|---|
| **Preserve routine registry data entry** | Retain the operational registry system as the source of data entry and build the FAIRification layer downstream. |
| **Support analytical reuse** | Transform source registry data into OMOP CDM v5.4. |
| **Preserve provenance** | Retain source variable names, source values, ontology annotations and mapping artefacts. |
| **Support semantic representation** | Generate RDF representations from selected harmonised data and metadata resources. |
| **Support SPARQL querying and validation** | Load RDF outputs into GraphDB named graphs to support predefined SPARQL queries, semantic validation and graph exploration. |
| **Support public aggregate visualisation** | Generate dashboard indicators from curated OMOP-derived public views. |
| **Support programmatic aggregate reuse** | Expose selected non-identifiable aggregate dashboard outputs and competency-question results through documented FastAPI endpoints and export functions. |
| **Support machine-readable aggregate reuse** | Represent selected dashboard and API outputs as public aggregate RDF resources. |
| **Support findability** | Publish registry and dataset metadata through ERDRI and the HemaFAIR FDP. |
| **Support reproducibility** | Use version-controlled mappings, scripted refresh procedures and validation checks. |
| **Support future extension** | Use registry-aware configuration and modular transformation logic. |

**Note:** *The table links FAIRification planning outputs and conceptual modelling requirements to concrete implementation choices within the HemaFAIR FAIRification workflow for the INHERENT use case.*

#### Notes on scope and limitations

This conceptual modelling document captures the design phase and implementation alignment of the HemaFAIR FAIRification workflow for the INHERENT use case. It should be treated as a planning and traceability artefact rather than as a complete ontology specification, a clinical analysis plan, or an epidemiological study protocol.

Several points should be noted:

1. Source-level annotations and mapping artefacts were manually curated and reviewed by clinical-domain, data and semantic-modelling specialists. Automated scripts consumed these artefacts during transformation, but the selection of ontology terms, OMOP concepts, unit mappings and predefined response-option mappings reflected expert-reviewed decisions rather than automatically inferred mappings.
2. Clinical and Registry Entries Semantic Model (CARE-SM)[27] and OMOP CDM were implemented as independent FAIRification branches with different intended scopes. CARE-SM provided a semantic representation of selected European Rare Disease Common Data Elements (EU RD CDEs)[15], whereas OMOP CDM provided the broader analytical harmonisation layer used for dashboard, aggregate FastAPI and CQ-oriented outputs.
3. The initial RDF materialisation used a controlled subset of the curated OMOP-derived layer to support reproducibility, semantic validation and SPARQL-based graph queries.
4. CYHAPR was implemented as an external INHERENT node, contributing to the registry-aware design and aggregate validation of the workflow. The manuscript remains centred on the INHERENT central platform FAIRification implementation, with CYHAPR used to demonstrate registry-aware reuse and controlled aggregate integration.
5. Additional semantic extensions may be added as ontology curation, CQ validation and registry requirements evolve.

### OMOP standardised vocabularies

This supplementary section documents the OMOP standardised vocabularies loaded into the HemaFAIR implementation. Recording the vocabulary inventory and version information supports reproducibility by documenting the terminology resources available during the registry-to-OMOP transformation. Vocabulary versions correspond to the Athena release used for this implementation and provide the reference terminology environment for concept mapping and harmonisation.

**Supplementary Table S9. OMOP standardised vocabulary inventory for the HemaFAIR implementation.**

| Vocabulary ID | Vocabulary name | Vocabulary version | Vocabulary reference | Category | Directly referenced by HemaFAIR mapping artefacts |
|---|---|---|---|---|---|
| ATC | WHO Anatomic Therapeutic Chemical Classification | ATC 2026-02-01 | http://www.whocc.no/atc_ddd_index/ | External/source vocabulary | No |
| CDM | OMOP Common Data Model | CDM v6.0.0 | https://github.com/OHDSI/CommonDataModel | OMOP supporting/internal | No |
| CMS Place of Service | Place of Service Codes for Professional Claims (CMS) | 2009-01-11 | http://www.cms.gov/Medicare/Medicare-Fee-for-Service-Payment/PhysicianFeeSched/downloads//Website_POS_database.pdf | External/source vocabulary | No |
| Cohort Type | OMOP Cohort Type | | OMOP generated | OMOP supporting/internal | No |
| Concept Class | OMOP Concept Class | | OMOP generated | OMOP supporting/internal | No |
| Condition Status | OMOP Condition Status | | OMOP generated | OMOP supporting/internal | No |
| Condition Type | OMOP Condition Occurrence Type | | OMOP generated | OMOP supporting/internal | No |

| Vocabulary ID | Vocabulary name | Vocabulary version | Vocabulary reference | Category | Directly referenced by HemaFAIR mapping artefacts |
|---|---|---|---|---|---|
| Cost | OMOP Cost | | OMOP generated | OMOP supporting/internal | No |
| Cost Type | OMOP Cost Type | | OMOP generated | OMOP supporting/internal | No |
| CPT4 | Current Procedural Terminology version 4 (AMA) | 2025 Release | http://www.nlm.nih.gov/research/umls/licensedcontent/umlsknowledgesources.html | External/source vocabulary | No |
| Death Type | OMOP Death Type | | OMOP generated | OMOP supporting/internal | No |
| Device Type | OMOP Device Type | | OMOP generated | OMOP supporting/internal | No |
| Domain | OMOP Domain | | OMOP generated | OMOP supporting/internal | No |
| Drug Type | OMOP Drug Exposure Type | | OMOP generated | OMOP supporting/internal | No |
| Episode | OMOP Episode | Episode 20201014 | OMOP generated | OMOP supporting/internal | No |
| Ethnicity | OMOP Ethnicity | Ethnicity 20250807 | OMOP generated | OMOP supporting/internal | No |
| Gender | OMOP Gender | | OMOP generated | OMOP supporting/internal | Yes |
| HCPCS | Healthcare Common Procedure Coding System (CMS) | 20260101 Alpha Numeric HCPCS File | http://www.nlm.nih.gov/research/umls/licensedcontent/umlsknowledgesources.html | External/source vocabulary | No |
| HPO | Human Phenotype Ontology | HPO v2025-11-24 | https://hpoo.jax.org | External/source vocabulary | Yes |

| Vocabulary ID | Vocabulary name | Vocabulary version | Vocabulary reference | Category | Directly referenced by HemaFAIR mapping artefacts |
|---|---|---|---|---|---|
| ICD10CM | International Classification of Diseases, Tenth Revision, Clinical Modification (NCHS) | ICD10CM FY2026 code descriptions | https://www.cdc.gov/nchs/icd/icd-10-cm.htm | External/source vocabulary | No |
| Korean Revenue Code | Korean Revenue Code (KNHIS) | | OMOP generated | OMOP supporting/internal | No |
| Language | OMOP Language | Language 20221030 | OMOP generated | OMOP supporting/internal | No |
| LOINC | Logical Observation Identifiers Names and Codes (Regenstrief Institute) | LOINC 2.80 | http://loinc.org/downloads/loinc | External/source vocabulary | Yes |
| Meas Type | OMOP Measurement Type | | OMOP generated | OMOP supporting/internal | No |
| Metadata | OMOP Metadata | | OMOP generated | OMOP supporting/internal | No |
| NCIt | NCI Thesaurus (National Cancer Institute) | NCIt 20220509 | http://evs.nci.nih.gov/ftp1/NCI_Thesaurus | External/source vocabulary | Yes |
| NDC | National Drug Code (FDA and manufacturers) | NDC 20260222 | http://www.nlm.nih.gov/research/umls/rxnorm/docs/rxnormfiles.html, http://www.fda.gov/downloads/Drugs/DevelopmentApprovalProcess/UCM070838.zip | External/source vocabulary | No |
| None | OMOP Standardized Vocabularies | v5.0 27-FEB-26 | OMOP generated | OMOP supporting/internal | No |
| Note Type | OMOP Note Type | | OMOP generated | OMOP supporting/internal | No |

| Vocabulary ID | Vocabulary name | Vocabulary version | Vocabulary reference | Category | Directly referenced by HemaFAIR mapping artefacts |
|---|---|---|---|---|---|
| NUCC | National Uniform Claim Committee Health Care Provider Taxonomy Code Set (NUCC) | NUCC 25.0 | http://www.nucc.org/index.php?option=com_content&view=article&id=107&Itemid=132 | External/source vocabulary | No |
| Observation Type | OMOP Observation Type | | OMOP generated | OMOP supporting/internal | No |
| Obs Period Type | OMOP Observation Period Type | | OMOP generated | OMOP supporting/internal | No |
| OMOP Extension | OMOP Extension (OHDSI) | OMOP Extension 20260202 | OMOP generated | OMOP supporting/internal | No |
| OMOP Genomic | OMOP Genomic vocabulary of known variants involved in disease | OMOP Genomic 20240216 | OMOP generated | OMOP supporting/internal | No |
| OSM | OpenStreetMap (OSMF) | OSM Release 2019-02-21 | https://www.openstreetmap.org/copyright/en, https://wambachers-osm.website/boundaries/ | External/source vocabulary | No |
| Plan | OMOP Health Plan | | OMOP generated | OMOP supporting/internal | No |
| Plan Stop Reason | OMOP Plan Stop Reason | | OMOP generated | OMOP supporting/internal | No |
| Procedure Type | OMOP Procedure Occurrence Type | | OMOP generated | OMOP supporting/internal | No |
| Race | Race and Ethnicity Code Set (USBC) | Version 1.0 | http://www.cdc.gov/nchs/data/dvs/Race_Ethnicity_CodeSet.pdf | External/source vocabulary | No |
| Relationship | OMOP Relationship | | OMOP generated | OMOP supporting/internal | No |

| Vocabulary ID | Vocabulary name | Vocabulary version | Vocabulary reference | Category | Directly referenced by HemaFAIR mapping artefacts |
|---|---|---|---|---|---|
| RxNorm | RxNorm (NLM) | RxNorm 20260105 | http://www.nlm.nih.gov/research/umls/rxnorm/docs/rxnormfiles.html | External/source vocabulary | Yes |
| RxNorm Extension | OMOP RxNorm Extension | RxNorm Extension 2026-01-14 | OMOP generated | OMOP supporting/internal | Yes |
| SNOMED | Systematic Nomenclature of Medicine - Clinical Terms (IHTSDO) | 2025-02-01 SNOMED CT International Edition; 2025-03-01 SNOMED CT US Edition; 2025-04-09 SNOMED CT UK Edition | http://www.nlm.nih.gov/research/umls/licensedcontent/umlsknowledgesources.html | External/source vocabulary | Yes |
| SOPT | Source of Payment Typology (PHDSC) | SOPT Version 9.2 | https://www.nahdo.org/sopt | External/source vocabulary | No |
| Sponsor | OMOP Sponsor | | OMOP generated | OMOP supporting/internal | No |
| Type Concept | OMOP Type Concept | Type Concept 20260202 | OMOP generated | OMOP supporting/internal | No |
| UB04 Point of Origin | UB04 Claim Source Inpatient Admission Code (CMS) | | https://www.resdac.org/cms-data/variables/Claim-Source-Inpatient-Admission-Code | External/source vocabulary | No |
| UB04 Pri Typ of Adm | UB04 Claim Inpatient Admission Type Code (CMS) | | https://www.resdac.org/cms-data/variables/Claim-Inpatient-Admission-Type-Code | External/source vocabulary | No |
| UB04 Pt dis status | UB04 Patient Discharge Status Code (CMS) | | https://www.resdac.org/cms-data/variables/patient-discharge-status-code | External/source vocabulary | No |

| Vocabulary ID | Vocabulary name | Vocabulary version | Vocabulary reference | Category | Directly referenced by HemaFAIR mapping artefacts |
|---|---|---|---|---|---|
| UB04 Typ bill | UB04 Type of Bill - Institutional (USHIK) | | https://ushik.ahrq.gov/ViewItemDetails?&system=apcd&itemKey=196987000 | External/source vocabulary | No |
| UCUM | Unified Code for Units of Measure (Regenstrief Institute) | Version 1.8.2 | http://aurora.regenstrief.org/~ucum/ucum.html#section-Alphabetic-Index | External/source vocabulary | Yes |
| UK Biobank | UK Biobank (UK Biobank) | version 2021-03-18 | https://biobank.ctsu.ox.ac.uk/showcase/schema.cgi; https://biobank.ctsu.ox.ac.uk/crystal/refer.cgi?id=141140 | External/source vocabulary | Yes |
| US Census | Census regions of the United States (USCB) | US Census 2017 Release | https://www.census.gov/geo/maps-data/data/tiger-cart-boundary.html | External/source vocabulary | No |
| Visit | OMOP Visit | Visit 20211216 | OMOP generated | OMOP supporting/internal | No |
| Visit Type | OMOP Visit Type | | OMOP generated | OMOP supporting/internal | No |
| Vocabulary | OMOP Vocabulary | | OMOP generated | OMOP supporting/internal | No |

***Note:*** *Vocabulary versions and references are those recorded in the OMOP* `vocabulary` *table following loading of Athena release v20260227, downloaded on 4 March 2026. Blank version fields reflect the corresponding values stored in the Athena vocabulary inventory for vocabularies that do not provide an explicit version identifier. The "Directly referenced by HemaFAIR mapping artefacts" column indicates whether a vocabulary is directly used in the curated HemaFAIR artefacts as a source ontology, target vocabulary, predefined response-option vocabulary or unit vocabulary. The version reported for the CDM vocabulary is the value recorded in the Athena vocabulary inventory and does not represent the HemaFAIR database schema version, which is OMOP CDM v5.4.*

### Representation of INHERENT registry data domains in the OMOP CDM

This supplementary section provides the domain-level mapping summary that underpins the HemaFAIR FAIRification workflow described in the main manuscript. It summarises how the major INHERENT eCRF domains were represented in the OMOP CDM and indicates the primary reuse scenario supported by each domain-level mapping.

**Supplementary Table S10** provides a high-level overview of domain representation. Some source domains are represented across multiple OMOP tables, depending on whether the source variable captures a clinical event, status, derived indicator, procedure, treatment exposure or provenance element. Field-level mappings, value maps and implementation artefacts are maintained in the project repository.

**Supplementary Table S10. Representation of INHERENT registry data domains in the OMOP CDM.**

| Data domain | Example source content | OMOP representation | Supported reuse scenario |
|---|---|---|---|
| **Patient demographics** | Sex, date of birth and age-related derived indicators | `person` | Cohort description, age and sex stratification |
| **Geography** | Country of birth, country of residence, centre country | `location`; `observation` for source-specific geographic context | Geographic stratification and dashboard filtering |
| **Treating or contributing centre** | Registry centre and healthcare provider institution | `care_site` | Centre-level aggregation and dashboard filtering |
| **Observation period and follow-up** | Baseline and annual update data periods | `observation_period` | Longitudinal anchoring and follow-up context |
| **Diagnosis** | Haemoglobinopathy diagnosis, diagnosis category and ORPHA- or SNOMED-linked codes | `condition_occurrence` | Diagnosis distribution and cohort definition |
| **Method of diagnosis** | Genetic testing, laboratory testing, clinical diagnosis and other diagnostic methods | `observation` | Diagnostic pathway characterisation |
| **Genotype and variants** | Gene identifiers, sequence variants, HGVS expressions and HGNC-related information | `measurement`; curated variant-lookup and provenance artefacts | Genotype stratification and genotype–phenotype analysis |
| **Laboratory measurements** | Haemoglobin, ferritin and other curated laboratory parameters | `measurement` | Longitudinal laboratory monitoring and biomarker analyses |

| Data domain | Example source content | OMOP representation | Supported reuse scenario |
|---|---|---|---|
| **Transfusion status** | Regular, occasional or no transfusion; transfusion-related status variables | `observation`; `procedure_occurrence` or treatment-related representation where applicable | Transfusion-status summaries and CQ1 validation |
| **Treatments and medications** | Iron chelation, hydroxyurea and other treatment-related variables | `drug_exposure`; `observation` for status or derived indicators | Treatment summaries and treatment-related filtering |
| **Procedures and surgical history** | Splenectomy and selected procedure-related events | `procedure_occurrence`; `observation` for status or history indicators | Procedure summaries and dashboard indicators |
| **Clinical complications and comorbidities** | Cardiac, endocrine, hepatic, renal, neurological, infectious and other manifestations | `condition_occurrence`; `observation` for status or contextual indicators | Complication monitoring and longitudinal analyses |
| **Consent and governance-related variables** | Consent categories, reuse permissions and data-sharing restrictions | `observation`; provenance and mapping artefacts | Governance-aware filtering and controlled reuse |
| **Mortality information** | Death status and death-related dates where available | `death`; `observation_period` | Survival and follow-up completeness analyses |
| **Source provenance** | Registry variable names, coded values, labels and ontology annotations | OMOP source-value fields and version-controlled mapping artefacts | Traceability, auditability and reproducible refresh |

**Note:** *The table summarises how major INHERENT eCRF domains were represented in the OMOP layer and their supported reuse scenarios. Some source domains are mapped to multiple OMOP tables depending on whether a source variable represents a clinical event, status, derived indicator, procedure, treatment exposure or provenance element.*

### SPARQL validation output for CQ1

This section presents an extended SPARQL validation output that supports the competency-question validation described in the main manuscript. **Supplementary Table S*11*** corresponds to CQ1, which assessed whether the OMOP-derived patient-level RDF graphs could support stratification by diagnosis group, country context, transfusion status and genotype availability. The table is provided as supplementary material because the full stratified output exceeds the size of the representative validation table included in the main text.

The output shows that the RDF graphs preserved the cross-domain links required to generate the CQ1 stratification. Counts below five were suppressed by the SPARQL query and are reported as "<5" to reduce disclosure risk. Differences in genotype availability across strata reflect the underlying registry data and should be interpreted as validation of graph connectivity rather than as epidemiological findings.

**Supplementary Table S11. Representative CQ1 SPARQL validation output from the OMOP-derived patient-level RDF graphs, stratified by diagnosis group, centre country, transfusion status and genotype availability.**

| Diagnosis group | Centre country | Transfusion status | Total patients | Patients with genotype information | Patients without genotype information |
|---|---|---|---|---|---|
| Alpha thalassemia (SNOMED:68913001) | Cyprus | No | 79 | 66 | 13 |
| Alpha thalassemia (SNOMED:68913001) | Cyprus | Unknown | 20 | 16 | <5 |
| Alpha thalassemia (SNOMED:68913001) | Cyprus | Yes (Occasional) | 33 | 31 | <5 |
| Alpha thalassemia (SNOMED:68913001) | Cyprus | Yes (Regular) | 14 | 14 | 0 |
| Alpha thalassemia (SNOMED:68913001) | Denmark | No | <5 | <5 | 0 |
| Alpha thalassemia (SNOMED:68913001) | Denmark | Unknown | <5 | <5 | 0 |
| Alpha thalassemia (SNOMED:68913001) | Denmark | Yes (Regular) | <5 | <5 | 0 |

| Diagnosis group | Centre country | Transfusion status | Total patients | Patients with genotype information | Patients without genotype information |
|---|---|---|---|---|---|
| Alpha thalassemia (SNOMED:68913001) | Greece | Unknown | <5 | <5 | 0 |
| Alpha thalassemia (SNOMED:68913001) | Greece | Yes (Regular) | <5 | <5 | 0 |
| Alpha thalassemia (SNOMED:68913001) | Malaysia | No | <5 | <5 | 0 |
| Alpha thalassemia (SNOMED:68913001) | Malaysia | Unknown | <5 | <5 | <5 |
| Alpha thalassemia (SNOMED:68913001) | Malaysia | Yes (Occasional) | <5 | <5 | 0 |
| Alpha thalassemia (SNOMED:68913001) | Malaysia | Yes (Regular) | 5 | 5 | 0 |
| Alpha thalassemia (SNOMED:68913001) | United States | Yes (Regular) | <5 | <5 | 0 |
| Beta thalassemia (SNOMED:65959000) | Cyprus | No | 20 | 20 | 0 |
| Beta thalassemia (SNOMED:65959000) | Cyprus | Unknown | <5 | <5 | <5 |
| Beta thalassemia (SNOMED:65959000) | Cyprus | Yes (Occasional) | 6 | 6 | 0 |
| Beta thalassemia (SNOMED:65959000) | Cyprus | Yes (Regular) | 493 | 434 | 59 |
| Beta thalassemia (SNOMED:65959000) | Denmark | No | <5 | <5 | 0 |
| Beta thalassemia (SNOMED:65959000) | Denmark | Unknown | <5 | <5 | 0 |
| Beta thalassemia (SNOMED:65959000) | Denmark | Yes (Occasional) | <5 | <5 | 0 |
| Beta thalassemia (SNOMED:65959000) | Denmark | Yes (Regular) | 10 | 10 | 0 |
| Beta thalassemia (SNOMED:65959000) | Greece | No | 9 | 9 | 0 |

| **Diagnosis group** | **Centre country** | **Transfusion status** | **Total patients** | **Patients with genotype information** | **Patients without genotype information** |
|---|---|---|---|---|---|
| Beta thalassemia (SNOMED:65959000) | Greece | Unknown | <5 | <5 | 0 |
| Beta thalassemia (SNOMED:65959000) | Greece | Yes (Occasional) | <5 | <5 | 0 |
| Beta thalassemia (SNOMED:65959000) | Greece | Yes (Regular) | 87 | 87 | 0 |
| Beta thalassemia (SNOMED:65959000) | Malaysia | No | <5 | <5 | 0 |
| Beta thalassemia (SNOMED:65959000) | Malaysia | Unknown | 19 | 19 | 0 |
| Beta thalassemia (SNOMED:65959000) | Malaysia | Yes (Occasional) | 14 | 14 | 0 |
| Beta thalassemia (SNOMED:65959000) | Malaysia | Yes (Regular) | 37 | 37 | 0 |
| Beta thalassemia (SNOMED:65959000) | Pakistan | Unknown | <5 | 0 | <5 |
| Beta thalassemia (SNOMED:65959000) | United States | Unknown | <5 | 0 | <5 |
| Sickle cell disease and related disorders (ORPHA:275752) | Angola | No | 10 | 0 | 10 |
| Sickle cell disease and related disorders (ORPHA:275752) | Angola | Unknown | 10 | 0 | 10 |
| Sickle cell disease and related disorders (ORPHA:275752) | Angola | Yes (Occasional) | 7 | 0 | 7 |
| Sickle cell disease and related disorders (ORPHA:275752) | Cyprus | No | 10 | 6 | <5 |
| Sickle cell disease and related disorders (ORPHA:275752) | Cyprus | Yes (Occasional) | 6 | <5 | <5 |

| Diagnosis group | Centre country | Transfusion status | Total patients | Patients with genotype information | Patients without genotype information |
|---|---|---|---|---|---|
| Sickle cell disease and related disorders (ORPHA:275752) | Cyprus | Yes (Regular) | <5 | 0 | <5 |
| Sickle cell disease and related disorders (ORPHA:275752) | Democratic Republic of the Congo | No | 57 | 0 | 57 |
| Sickle cell disease and related disorders (ORPHA:275752) | Democratic Republic of the Congo | Unknown | 19 | 0 | 19 |
| Sickle cell disease and related disorders (ORPHA:275752) | Democratic Republic of the Congo | Yes (Occasional) | 46 | 0 | 46 |
| Sickle cell disease and related disorders (ORPHA:275752) | Denmark | No | <5 | <5 | 0 |
| Sickle cell disease and related disorders (ORPHA:275752) | Denmark | Unknown | 11 | 11 | 0 |
| Sickle cell disease and related disorders (ORPHA:275752) | Denmark | Yes (Occasional) | <5 | <5 | 0 |
| Sickle cell disease and related disorders (ORPHA:275752) | Denmark | Yes (Regular) | <5 | <5 | 0 |
| Sickle cell disease and related disorders (ORPHA:275752) | Greece | No | 7 | 7 | 0 |
| Sickle cell disease and related disorders (ORPHA:275752) | Greece | Unknown | <5 | <5 | 0 |
| Sickle cell disease and related disorders (ORPHA:275752) | Greece | Yes (Occasional) | <5 | <5 | 0 |
| Sickle cell disease and related disorders (ORPHA:275752) | Greece | Yes (Regular) | 5 | 5 | 0 |

| Diagnosis group | Centre country | Transfusion status | Total patients | Patients with genotype information | Patients without genotype information |
|---|---|---|---|---|---|
| Sickle cell disease and related disorders (ORPHA:275752) | Malaysia | No | <5 | <5 | 0 |
| Sickle cell disease and related disorders (ORPHA:275752) | Malaysia | Unknown | <5 | <5 | 0 |
| Sickle cell disease and related disorders (ORPHA:275752) | Malaysia | Yes (Occasional) | <5 | <5 | 0 |
| Sickle cell disease and related disorders (ORPHA:275752) | Nigeria | No | 23 | 14 | 9 |
| Sickle cell disease and related disorders (ORPHA:275752) | Nigeria | Unknown | 152 | 15 | 137 |
| Sickle cell disease and related disorders (ORPHA:275752) | Nigeria | Yes (Occasional) | 21 | <5 | 18 |
| Sickle cell disease and related disorders (ORPHA:275752) | Nigeria | Yes (Regular) | <5 | 0 | <5 |
| Sickle cell disease and related disorders (ORPHA:275752) | United States | No | 26 | 26 | 0 |
| Sickle cell disease and related disorders (ORPHA:275752) | United States | Unknown | 61 | 54 | 7 |
| Sickle cell disease and related disorders (ORPHA:275752) | United States | Yes (Occasional) | <5 | <5 | 0 |
| Sickle cell disease and related disorders (ORPHA:275752) | United States | Yes (Regular) | <5 | <5 | <5 |

***Note:*** *The query was executed across the INHERENT and CYHAPR named graphs containing OMOP-derived patient-level RDF. The output reports patient counts stratified by diagnosis group, centre country and transfusion status, together with the numbers of patients with and without genotype information in each stratum. Counts below five were suppressed independently within each reported count field and are shown as "<5" to reduce disclosure risk. The table presents a representative validation snapshot generated on 30 June 2026.*

### References


1. Kountouris, P. et al. The International Hemoglobinopathy Research Network (INHERENT): An international initiative to study the role of genetic modifiers in hemoglobinopathies. *Am. J. Hematol.* 96, E416–E420 (2021). doi:10.1002/ajh.26323.
2. Kountouris, P. et al. Pilot of the International Hemoglobinopathy Research Network for a multiethnic genome-wide association study. *Blood Glob. Hematol.* 2, 100098 (2026). doi:10.1016/j.bglo.2026.100098.
3. Bernabé, C. *et al.* GO-Plan: A goal-oriented method for FAIRification planning. *Information Services and Use* **45**, 100–124 (2025).
4. Voss, E. A. *et al.* Feasibility and utility of applications of the common data model to multiple, disparate observational health databases. *J Am Med Inform Assoc* **22**, 553–564 (2015).
5. OMOP CDM v5.4. https://ohdsi.github.io/CommonDataModel/cdm54.html.
6. RDF - Semantic Web Standards. https://www.w3.org/RDF/.
7. What is GraphDB? — GraphDB 11.3 documentation. https://graphdb.ontotext.com/documentation/11.3/.
8. INHERENT. INHERENT Dashboard. *INHERENT Public Dashboard* https://dashboard.inherentnetwork.org/.
9. HemaFAIR - INHERENT. INHERENT Dashboard API - Swagger UI. *INHERENT* https://dashboard.inherentnetwork.org/inherent-dashboard-api/docs.
10. FAIR Data Point. INHERENT. https://w3id.org/HemaFAIR/fdp.
11. Wilkinson, M. D. *et al.* The FAIR Guiding Principles for scientific data management and stewardship. *Sci Data* **3**, 160018 (2016).
12. HemaFAIR. https://hemafairproject.eu/ (2024).

13. Harris, P. A. *et al.* The REDCap consortium: Building an international community of software platform partners. *Journal of Biomedical Informatics* **95**, 103208 (2019).
14. Ramírez, S. FastAPI. https://fastapi.tiangolo.com.
15. European Platform on Rare Disease EU-RD. https://eu-rd-platform.jrc.ec.europa.eu.
16. daSilvaSantos, L., Burger, K., Kaliyaperumal, R. & Wilkinson, M. FAIR Data Point: A FAIR-Oriented Approach for Metadata Publication. *Data Intell.* 5, 163–183 (2023).
17. Archer, N. et al. Multi-Ethnic, Resource-Adaptable Case Report Form (CRF) by the International Hemoglobinopathy Research Network (INHERENT). (2026). doi:10.5281/zenodo.21260336.
18. Weinreich, S. S., Mangon, R., Sikkens, J. J., Teeuw, M. E. & Cornel, M. C. [Orphanet: a European database for rare diseases]. *Ned Tijdschr Geneeskd* **152**, 518–519 (2008).
19. Stearns, M. Q., Price, C., Spackman, K. A. & Wang, A. Y. SNOMED clinical terms: overview of the development process and project status. *Proc AMIA Symp* 662–666 (2001).
20. Köhler, S. *et al.* The Human Phenotype Ontology in 2021. *Nucleic Acids Research* **49**, D1207–D1217 (2021).
21. McDonald, C. J. *et al.* LOINC, a Universal Standard for Identifying Laboratory Observations: A 5-Year Update. *Clinical Chemistry* **49**, 624–633 (2003).
22. Povey, S. *et al.* The HUGO Gene Nomenclature Committee (HGNC). *Hum Genet* **109**, 678–680 (2001).
23. Hart, R. K. *et al.* HGVS Nomenclature 2024: improvements to community engagement, usability, and computability. *Genome Med* **16**, 149 (2024).
24. RxNorm Overview. https://www.nlm.nih.gov/research/umls/rxnorm/overview.html.

25. ERDRI.dor - International Hemoglobinopathy Research Network. *European Rare Disease Registry Infrastructure* https://eu-rd-platform.jrc.ec.europa.eu/erdridor/register/name/INHERENT.

26. ERDRI.mdr - International Hemoglobinopathy Research Network. *European Rare Disease Registry Infrastructure* https://eu-rd-platform.jrc.ec.europa.eu/mdr/detail/INHERENT.

27. Kaliyaperumal, R. *et al.* Semantic modelling of common data elements for rare disease registries, and a prototype workflow for their deployment over registry data. *Journal of Biomedical Semantics* **13**, 9 (2022).
